# Globally altered epigenetic landscape and lagging osteogenic differentiation in H3.3-G34W-mutant giant cell tumor of bone

**DOI:** 10.1101/2020.05.26.20089888

**Authors:** Pavlo Lutsik, Annika Baude, Daniela Mancarella, Simin Öz, Alexander Kühn, Reka Toth, Joschka Hey, Umut H. Toprak, Jinyeong Lim, Viet Ha Nguyen, Chao Jiang, Anand Mayakonda, Mark Hartmann, Felix Rosemann, Kersten Breuer, Dominik Vonficht, Florian Grünschläger, Suman Lee, Maren Kirstin Schuhmacher, Denis Kusevic, Anna Jauch, Dieter Weichenhan, Jozef Zustin, Matthias Schlesner, Simon Haas, Joo Hyun Park, Yoon Jung Park, Udo Oppermann, Albert Jeltsch, Florian Haller, Jörg Fellenberg, Anders M. Lindroth, Christoph Plass

**Author notes:** These authors contributed equally (Pavlo Lutsik, Annika Baude, Daniela Mancarella, Simin Öz, Anders M. Lindroth). **Corresponding authors** Christoph Plass, Anders Lindroth.

## Abstract

The neoplastic stromal cells of giant cell tumor of bone (GCTB) carry a mutation in *H3F3A*, leading to a mutant histone variant, H3.3-G34W, as a sole recurrent genetic alteration. We show that in patient-derived stromal cells H3.3-G34W is incorporated into the chromatin and associates with massive epigenetic alterations on the DNA methylation, chromatin accessibility and histone modification level that can be partially recapitulated in an orthogonal cell line system by the introduction of H3.3-G34W. These epigenetic alterations affect mainly heterochromatic and bivalent regions and provide possible explanations for the genomic instability as well as the osteolytic phenotype of GCTB. The mutation occurs in differentiating mesenchymal stem cells and associates with an impaired osteogenic differentiation. We propose that the observed epigenetic alterations reflect distinct differentiation stages of H3.3 WT and H3.3 MUT stromal cells and add to H3.3-G34W associated changes.

**Important abbreviations:** H3.3-G34W, mutated histone variant; H3.3 MUT, stromal cells expressing H3.3-G34W; H3.3 WT, stromal cells expressing wildtype H3.3

## Introduction

The discovery of mutated histone genes in aggressive cancers raised a lot of interest in the cancer research community due to their ability to globally alter the epigenomic landscape (1). A frequently mutated histone is the non-canonical histone variant H3.3 (2,3). In contrast to canonical H3.1 and H3.2, the incorporation of histone variant H3.3 is replication-independent and its turnover occurs throughout the cell cycle (4). Deposition occurs either via the HIRA chaperone complex at sites of gene activation or through ATRX-DAXX into heterochromatic regions (5) and the silent allele of imprinted genes (6). Mouse embryonic stem cells require H3.3 for correct establishment of H3K27me3 patterns at bivalent promoters of developmentally regulated genes (7,8). While two human genes, *H3F3A* and *H3F3B*, encode for an identical H3.3 protein, oncogenic mutations occur gene-specifically in different tumor types. Lysine-27-to-methionine (H3.3-K27M) and glycine-to-arginine or valine substitution (H3.3-G34R/V) in pediatric gliomas (9), as well as glycine-34-to-tryptophan or leucine (H3.3-G34W/L) substitutions in giant cell tumor of bone (GCTB) (10) have been described, all due to mutations in *H3F3A*. For *H3F3B*, mutations leading to a lysine -36-to-methionine (H3.3-K36M) substitution were reported in chondroblastomas (10,11). The molecular consequence of the H3.3-K27M mutation is a global loss of the repressive chromatin mark H3K27me3 through inactivation of the PRC2 complex (12-14). Similarly, the H3.3-K36M mutation suppresses the deposition of the H3K36me3 mark by interference with histone methyltransferases NSD2 and SETD2 (11,15). Recent findings suggested reduced levels of H3K36me3 and increased levels of H3K27me3 *in cis* in HeLa cells overexpressing H3.3-G34W (16). However, the detailed effects of this mutant histone variant on the epigenome are yet to be determined and to be analyzed in patients. GCTB, where this mutation was shown as the sole alteration, offers a unique system to study these effects in primary patient material.

GCTB is a rare locally aggressive bone neoplasm, which typically affects the meta-epiphyseal regions of long bones in young adults (17). These tumors consist of three major cell types: stromal cells originating from mesenchymal stem cells (MSC), multinuclear giant cells and mononuclear histiocytic cells (18). The GCTB stromal cells show incidence of H3.3-G34W in more than 90% of cases and display markers of both MSC and pre-osteoblast cell populations (10,19). The neoplastic stromal cell population secretes high levels of the receptor activator of NF-κB ligand (RANKL) and reduced levels of its decoy receptor, osteoprotegerin (OPG), thereby attracting and activating surrounding monocytes. Upon activation, the recruited monocytes fuse to form multinucleated giant cells, which resemble osteoclasts and lead to massive bone destruction (17).

Here we investigate the effects of H3.3-G34W on global epigenomic patterns in patient samples from four different centers. We find epigenetic distortions that contribute to the phenotypes of GCTB, stochastic genomic instability and increased osteolysis. Furthermore, we demonstrate that neoplastic and non-neoplastic GCTB stromal cells represent distinct stages of osteogenic differentiation. Differentiation-related epigenetic differences add to the overall picture of H3.3-G34W-associated global epigenetic alterations, whereas the differentiation delay is potentially driven by the direct effects of H3.3-G34W. Our findings collectively suggest that the single alteration of G34W induces epigenomic changes with implications for the development of stromal cells and the tumorigenic process.

## Results

### H3K36 methylation levels are not changed in H3.3-G34W-expressing stromal cells from GCTB patients

Recent biochemical studies have shown that G34 substitutions in H3.3, including G34W, inhibit the activity of the histone methyltransferase SETD2, which is responsible for H3K36me3, *in cis* (16). We verified these effects in HEK293 cells stably overexpressing H3.3-G34W or wild-type H3.3 as a control and confirmed *in cis* effects on H3.3-G34W K36 trimethylation levels. We did not observe any *in trans* effects on endogenous H3 modifications as found for other mutant histones such as H3-K36M (**Figure S1a**).

To specifically test if these biochemical findings apply to patient samples, we obtained access to GCTB biopsies from four different cohorts (**Table S1**). For the initial characterization of 30 GCTB samples (**Table S1**), we performed immunohistochemical analysis with a H3.3-G34W-specific antibody. Positive staining was observed and validated in 29 of 30 cases (**Fig. 1a** and **Fig. S1b**). The H3.3-G34W-negative case (unified patient identifier, UPI-13) carried a *H3F3A* (c.103_104GG>TT) mutation encoding a H3.3-G34L substitution that has already been described for GCTB (10) (**Fig. 1b, S1c**). We established both, neoplastic, H3.3 G34W-expressing (H3.3 MUT) and non-neoplastic, H3.3-G34W negative (H3.3 WT) stromal cell lines (**Fig. 1b; Table S1**), from primary tissue. Cell type differences were ruled out by flow cytometric analyses which revealed a high expression of MSC markers and low expression of hematopoietic markers in both H3.3 WT and H3.3 MUT cell lines, suggesting that both are of mesenchymal origin (**Fig. S1d**). In total, we collected 96 tissue samples from 95 different GCTB patients from four different cohorts (**Table S1**). We were able to establish 26 stromal cell lines from 24 different GCTB patients from two cohorts (**Fig. 1c**). In addition to H3.3 WT cells, we analyzed bone marrow-derived primary MSCs from non-GCTB patients from here on referred to as nontumoral stromal cells (nt-SC) (**Table S1**). We verified the mutational status of the cells using Sanger resequencing, ultra-deep resequencing on the MiSeq platform or whole-genome sequencing (**Table S1, Fig. 1b, S1c, S1e**). In addition to the common mutation leading to the G34W alteration in H3.3, whole-genome sequencing of seven patient-derived primary H3.3 MUT cell lines (**Table S1**) did not reveal any recurrent genetic alterations (**Fig. S1e**). In contrast to other malignancies carrying *H3F3A* mutations, as for example pediatric glioblastoma with co-occurring mutations in TP53 and ATRX (9,20), and to other bone tumors, e.g. osteosarcoma, GCTB showed an extremely low overall mutation frequency for H3.3 MUT and H3.3 WT cells (**Fig. 1d, Fig. S1e**). This places GCTB in a unique position to uncover epigenomic alterations linked to H3.3-G34W.

**Figure 1:**
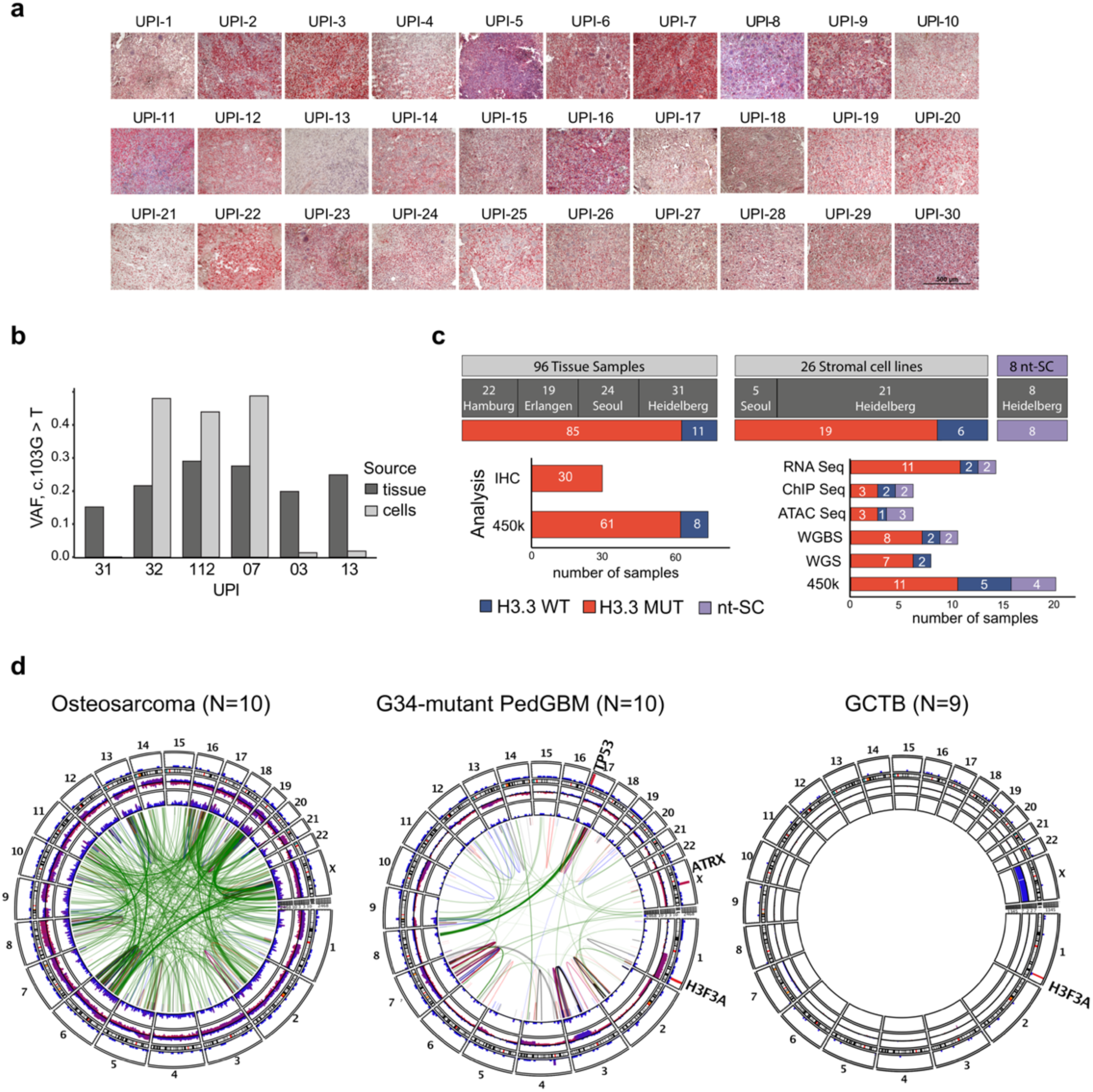
Initial characterization of GCTB patient samples. **a**. Immunohistochemical staining of primary GCTB tumor resections with a H3.3-G34W-specific antibody (Active Motif) (red). UPI, unified patient identifier. **b**. Quantification of the mutation at position 103 in the *H3F3A* gene (c.103G>T) leading to the H3.3 G34W substitution in tumor resections and derived stromal cell lines using deep targeted resequencing. VAF, variant allele frequency. **c**. Overview of GCTB tissues and derived stromal cell lines analyzed within this paper. **d**. Circos plot of recurrent structural variants in osteosarcoma, H3.3-G34R-baring pediatric glioblastoma (PedGBM) and GCTB cohorts based on whole genome sequencing data. Green lines represent translocations, blue lines deletions, red lines duplications, and black lines inversions. The variant recurrences are represented by bar plots. The outermost layer represents functional small variants (SNVs and small indels). The middle layer represents copy number variations. The innermost layer represents structural variations. All layers are normalized to the compared cohort size. Osteosarcoma cohort was sub-sampled at random to the size of the other two cohorts.

To understand how H3.3-G34W exerts its function in tumor cells, we analyzed protein fractions by Western blot and found H3.3-G34W incorporated into chromatin (**Fig. S2a**). To analyze *in cis* effects of H3.3-G34W in patient derived cells, we used a specific and verified antibody to map H3.3-G34W and identify sites of H3.3-G34W enrichment in two independent patient cell lines. We identified high-confidence genomic regions showing H3.3-G34W enrichment (**Fig. S2b, c, Table S2**) and profiled H3K36me3 along with several other histone modifications using ChIP-mentation. Changes of H3K36me3 or H3K27me3 between H3.3 WT and H3.3 MUT stromal cells at sites of H3.3-G34W enrichment were minute and did not recapitulate the *in cis* results observed in HEK293 cells (**Fig. S2b, c**). Furthermore, using Western blot analysis of whole cell lysates we did not reveal any changes in the total amount of K36 methylated histone H3 indicating that the G34W substitution does not affect methylation of K36 on other histones lacking the mutation. (**Fig. S2d**).

### The epigenome of GCTB H3.3 MUT cells is globally altered

We hypothesized that H3.3-G34W might exert its effects through other unidentified epigenetic mechanisms and screened H3.3 MUT and H3.3 WT cells, as well as a large number of biopsies of the four GCTB cohorts, using HumanMethylation450 arrays (89 samples in total). We observed full-range DNA methylation changes of a large number of CpG sites (**Fig. 2a**). Dimensionality reduction and clustering analysis of the methylation data showed the most pronounced changes between primary patient-derived H3.3 MUT and H3.3 WT cell lines as compared to GCTB biopsies. Most of the primary tumor samples had an intermediate methylome identifying them as mixtures of normal and tumor cells (**Fig. 2a, b**), as also confirmed by deep resequencing of the *H3F3A* gene locus of tissue-derived DNA showing lower variant allele frequencies than theoretical 0.5 for a heterozygous mutation (**Fig. 1b**). This observation prompted us to restrict all our subsequent analyses to H3.3 MUT and H3.3 WT cells in order to obtain a clean view of H3.3-G34W-associated epigenetic alterations in pure cell populations. To further a detailed analysis of the epigenome, we profiled DNA methylation at single CpG resolution using whole genome bisulfite sequencing (WGBS), analyzed chromatin accessibility with the assay for transposase-accessible chromatin using sequencing (ATAC-seq) and analyzed global distribution of several histone marks (H3K4me1, H3K4me3, H3K9me3, H3K27me3, H3K27ac, H3K36me3) to investigate their potential redistribution. Global hierarchical clustering proved the H3.3-G34W substitution to be the major determinant of variability in DNA methylation, chromatin accessibility and posttranslational histone modifications (H3K27me3, H3K27ac, H3K4me1) with H3.3 MUT and H3.3 WT groups forming separate clusters (**Fig. 2c, Fig. S2e**). In contrast, other modifications (H3K36me3, H3K4me3) did not yield in H3.3 MUT- and H3.3 WT-separating clusters. Since nt-SCs clustered with H3.3 WT cells, we combined all H3.3 wild type cells into a single control group (H3.3 WT) for further analysis.

**Figure 2:**
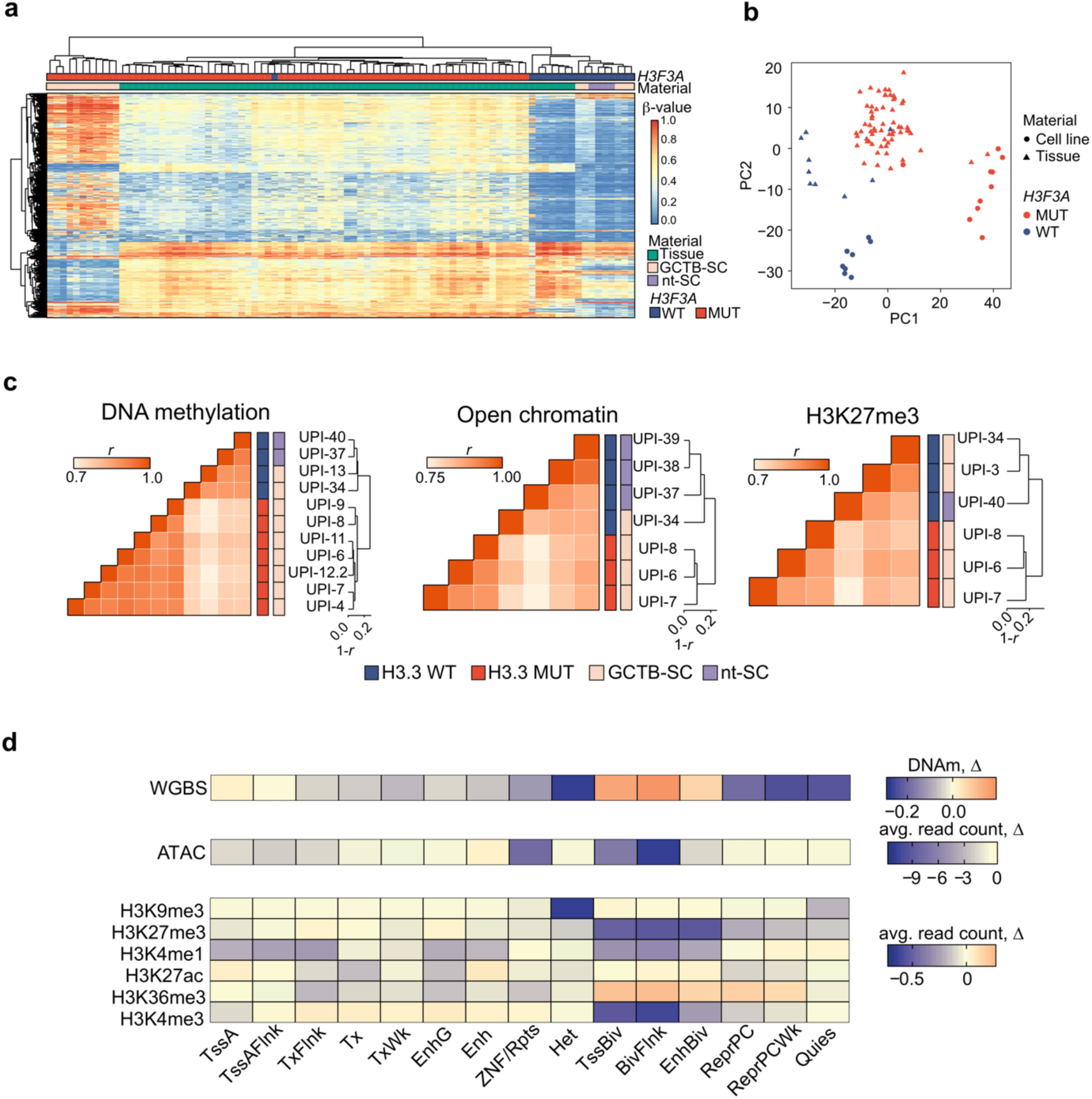
Genome-wide epigenetic distortion in H3.3 MUT cells. **a**. HumanMethylation450 profiles of GCTB tumor tissue samples, GCTB stromal cells and healthy nontumoral stromal cells (nt-SC). Heatmap displays 10,000 CpG sites with highest S.D. across all samples. Agglomerative, hierarchical clustering of rows (CpGs) and columns (samples) was performed with average-linkage based on Euclidean distance metric. **b**. Principal component analysis of HumanMethylation450 array-based DNA methylation profiles of GCTB tumor tissue, GCTB stromal cells and nontumoral stromal cells (nt-SC). **c**. Hierarchical clustering with correlation distance of DNA methylation (left), open chromatin (middle) and H3K27me3 profiles (right) in H3.3 WT (blue) and H3.3 MUT (red) cells. Heatmaps represent pairwise Pearson correlation coefficients (*r*) between respective modification profiles of two cell lines. Dendrograms were obtained with agglomerative hierarchical clustering with 1-r distance and average linkage. Heatmap color codes represent *H3F3A* mutational status (inner), and the cell type (outer), the common legend for all heatmaps is given at the bottom. nt-SC-nontumoral stromal cells. UPI, Unified patient identifier. **d**. Stratification of epigenetic differences using MSC-specific chromatin states as defined by ChromHMM. For DNA methylation (WGBS, “DNAm”) difference between average methylation levels of all CpGs falling into corresponding states in H3.3 WT and H3.3 MUT cells is presented. For ATAC and ChIP-seq of histone modifications, the difference between average normalized read counts over all windows of a state is shown. Mnemonics for the ChromHMM states are defined by Roadmap (TssA, active TSS; TssFlnk, active TSS flanking regions; Tx, transcribed regions; TxFlnk, transcription franking regions; TxWk, weakly transcribed regions; Enh, enhancers; EnhG, genic enhancers, ZNF/Rpts; zinc finger genes and repeats; TssBiv, bivalent TSS; BivFlnk, flanking bivalent regions; EnhBiv, bivalent enhancer; ReprPC, Polycomb repressed; ReprPCWk, weak Polycomb repressed; Quies, quiescent).

In order to identify loci with more pronounced epigenetic changes, we used a genome-wide approach to stratify the observed differences from all epigenetic layers using the ENCODE functional annotation of the MSC epigenome (21). We observed profound differences in several groups of chromatin states including heterochromatic and Polycomb-repressed states as well as bivalent domains (**Fig. 2d**). Heterochromatic regions were noticeably hypomethylated while bivalent domains accumulated a whole range of alterations, including gain of DNA methylation, loss of chromatin accessibility, as well as loss of several histone modifications (H3K27me3, H3K4me3 etc). Collectively, we conclude that the epigenome of H3.3 MUT cells shows significant and reproducible differences to that of H3.3 WT stromal cells.

### DNA hypomethylation in genomic megabase-scale domains indicates defects of heterochromatin potentially contributing to stochastic genomic rearrangements

We first followed up changes associated with loss of DNA methylation. In contrast to the homogeneity of DNA methylation profiles observed within each of the two groups (**Fig. S3a**), DNA methylation was non-uniformly altered between H3.3 WT and H3.3 MUT cells across the entire genome (**Fig. 3a**). The majority of significantly changed loci showed profound reduction of DNA methylation in H3.3 MUT cells, while a minority was hyper-methylated (**Fig.3a, Fig. S3b**). Overall, H3.3 MUT cells exhibited a 20% genome-wide reduction of DNA methylation (**Fig.3b, Fig. S3c**). To systematically characterize the abundant DNA methylation changes, we segmented the genome into large methylation domains (LMD I to IV) with sizes >20kb (**Fig. S3d, Table S2**) using a combination of breakpoint analysis and clustering (see Methods for details). The majority of regions (predominantly LMDs III and IV) matched the criteria for partially methylated domains i.e. megabase-scale domains of predominantly repressive chromatin with low gene density (22). Each LMD had a distinct level of DNA methylation, a discrete pattern of histone marks and gene density, suggesting different functional roles (**Fig. 3c-e**). For instance, LMD III showed enrichment of H3K27me3, while LMD IV was associated with H3K9me3, suggesting an association with facultative and constitutive heterochromatin (**Fig. 3e**). Furthermore, LMD III domains were often detected as flanking to LMD IV. LMDs I to IV showed decreasing average methylation levels starting from over 0.75 down to less than 0.25 (**Fig. 3c**). Lowly methylated LMDs III and IV which could be characterized as heterochromatic (**Fig. 3e**) showed the most pronounced demethylation comparing H3.3 MUT cells to H3.3 WT cells. We concluded that global epigenetic alterations in GCTB are non-uniformly distributed along the genome, and that the most pronounced global changes take place in large-scale domains associated with facultative and constitutive heterochromatin, roughly corresponding to LMDs III and IV.

**Figure 3:**
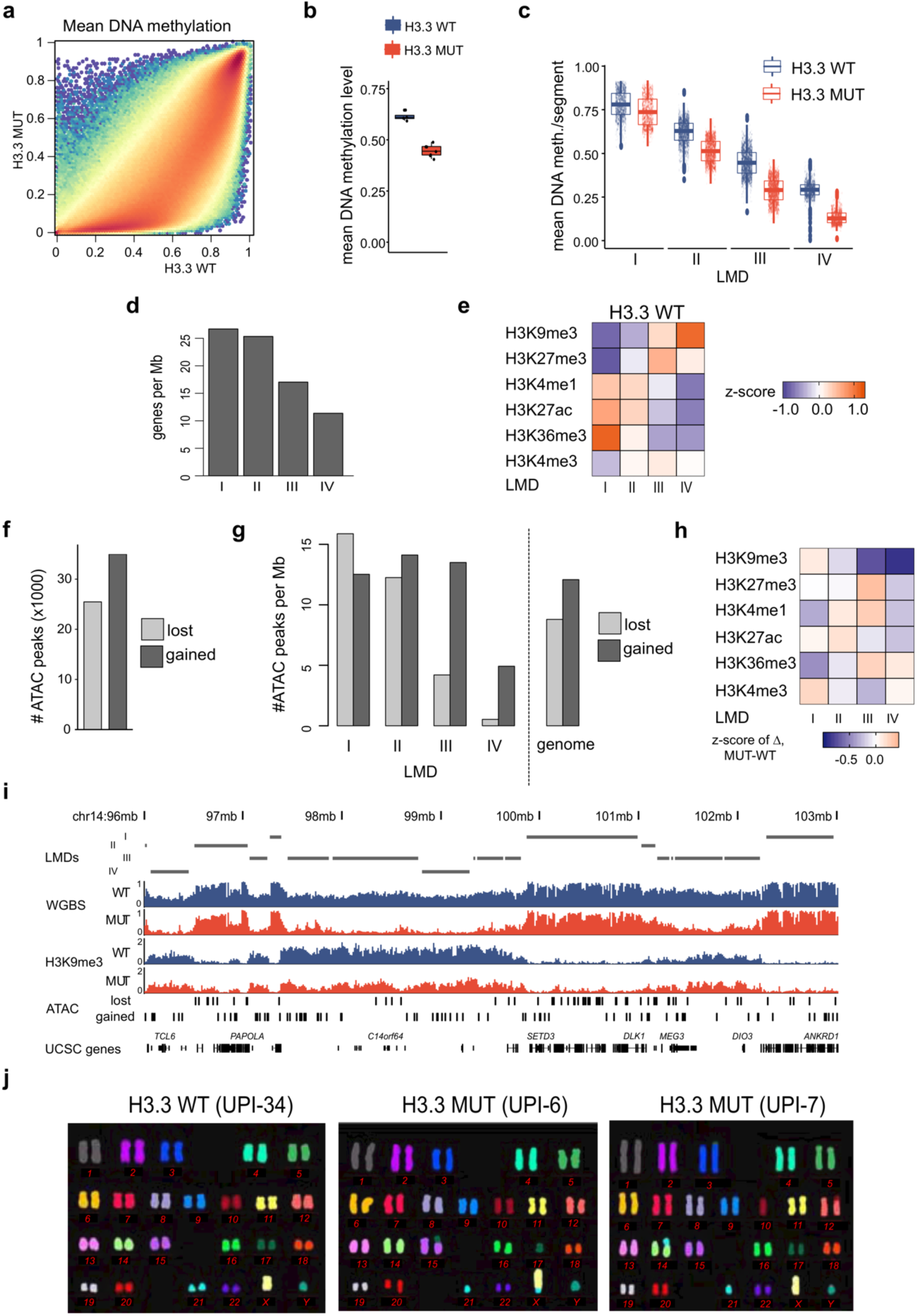
Heterochromatin defects in H3.3 MUT stromal cells. (on the next page). **a**. Binned scatterplot of DNA methylation profiles from WGBS of H3.3 WT cells versus H3.3 MUT cells. Hexagon color represents binned density gradient of 1 (blue), 1000 (yellow), and 10^6^ (red) points. **b**. Genome-wide mean level of DNA methylation in H3.3 WT (blue) and H3.3 MUT (red) cells. Points represent individual patients, box-plot bar represents the mean, box the inter-quartile range, whiskers extend from the smallest to the largest value within 1.5 IQR from the lower and upper edges of the box, respectively. The difference is statistically significant *(p*=1.5・10^-5^, two-sample t-test). **c**. Methylation level distribution at large methylation domains (LMDs) in H3.3 WT (blue) and H3.3 MUT (red) cells. Each point represents a single LMD segment. **d**. Mean gene density of LMDs. RefSeq genes were used and for each segment the number of overlapping genes was divided by its length in Mb. **e**. Normalized levels of histone modifications at LMDs in H3.3 WT cells. Heatmap represents z-scores of average signals across all segments falling into the LMDs calculated using overall mean level and standard deviation across all LMD segments. **f**. Absolute numbers of ATAC-seq peaks with differential chromatin accessibility between H3.3 WT cells and H3.3 MUT cells. “lost” and “gained” refer to peaks showing a decrease (disappearance) or increase (emergence) in H3.3 MUT cells, respectively. **g**. Stratification of differential chromatin accessibility between H3.3 WT and H3.3 MUT cells by overlap with LMDs. Labels “lost” and “gained” differential ATAC peaks as in (**f**); “genome”, distribution of ATAC peaks across the complete genome. **h**. Changes of histone modifications between H3.3 WT and H3.3 MUT stromal cells at LMDs. Heatmap represents difference of average read counts, Z-score-normalized with respect to zero. **i**. Genomic browser view of a representative 7Mbp region on chromosome 14 (positions 96,000,000–103,000,000) for whole-genome bisulfite sequencing, H3K9me3 ChIP-seq and ATAC-seq. Tracks are shown for H3.3 WT (blue) and H3.3 MUT (red) cells. Bottom panel indicates gene locations based on the UCSC default gene track. “lost” and “gained” differential ATAC-seq peaks as in (**f**). **j**. Visualization of karyotypes using m-FISH in H3.3 WT and H3.3 MUT cells.

We next sought to associate hypomethylation with other epigenetic alterations. Consistent with the global loss of DNA methylation, we observed increased chromatin accessibility in the H3.3 MUT cells with approximately 1.5-times more gained than lost differential ATAC-seq peaks (**Fig. 3f**). ATAC peaks gained in H3.3 MUT cells were overrepresented in the LMDs III and IV (**Fig. 3g**) indicating a more open state in respective genomic segments. These LMDs also showed strong reduction in the heterochromatic histone mark H3K9me3 (**Fig. 3h**). Examples of changes in different LMDs can be seen in **Fig. 3i**. The ATAC peaks gained in H3.3 MUT showed a significant overlap with repetitive elements such as LINE, SINE and LTR elements which are normally silenced by DNA methylation (**Fig. S3e, Table S3**). Looking at other known repetitive regions, such as centromeres and telomers, we found them to be affected by the genome-wide hypomethylation (**Fig. S3f**). m-FISH analysis found H3.3 WT cells to have a normal karyotype, while H3.3 MUT cells in contrast displayed different, non-recurrent centromeric fusions which could be a potential consequence of the heterochromatin defects described above (**Fig. 3j**). We conclude that H3.3-G34W associates with heterochromatic defects that potentially contribute to a genomic instability previously described as characteristic for GCTB (23).

### Isogenic cells expressing H3.3-G34W recapitulate DNA methylation changes found in H3.3 MUT stromal cells

As H3.3 MUT stromal cells showed the strongest epigenetic differences to H3.3 WT stromal cells on the DNA methylation level, we expect DNA methylation to play a major role in stromal cell transformation leading to GCTB. To verify the observed epigenetic changes to be dependent on H3.3-G34W expression, we aimed to recapitulate the findings in an unrelated cell line system. To this end, we introduced the H3.3-G34W encoding mutation in HeLa cells by targeting the endogenous *H3F3A* locus as earlier described (**Fig. S4a**) (24). Individual iso-H3.3- WT and iso-H3.3-G34W clones were isolated of which four iso-H3.3-WT and iso-H3.3-G34W monoclonal lines and one parental HeLa cell line were subject to HumanMethylationEPIC DNA methylation analysis (**Fig. 4a**). The iso-H3.3-G34W samples showed similar alterations as found when comparing H3.3 MUT and H3.3 WT stromal cells of GCTB (**Fig. 4a-e**). Principal component analysis of the most variable methylation probes clustered iso-H3.3-WT clones together while iso-H3.3-G34W clones dispersed off, indicating changes in DNA methylation in iso-H3.3-G34W but not WT isogenic cell lines (**Fig. 4a**). Similar to primary GCTB cells, the largest principal component (PC1, 31% of variance explained) captured widespread hypomethylation in iso-H3.3-G34W accompanied by focal hypermethylation events (Fig. 4a). Clustering analysis further enforced the distinct difference between iso-H3.3- WT and G34W clones (**Fig. 4b**). A scatter plot of all probes indicated that iso-H3.3-G34W cells showed predominantly hypomethylation like H3.3 MUT cells from GCTB (**Fig. 4c**). We found 9,047 differentially methylated probes (Δ ☐-value>0.2 and FDR<0.05) of which 5,688 (63%) were hypomethylated in iso-H3.3-G34W cells (**Fig. 4d**). CpG sites that lost methylation were more frequently localized to intergenic regions and promoter-proximal exons, while hypermethylated ones were in addition moderately enriched at promoters and depleted at exons (**Fig. 4e**). To a large extent, this recapitulated the situation in primary GCTB stromal cells and confirmed that the changes in DNA methylation are associated with H3.3-G34W. As a specific example, the *RANKL* locus showed hypomethylation at exon 3, a potential alternative promoter element, (**Fig. 4f**) which is in line with the methylation differences observed between H3.3 WT and H3.3 MUT stromal cells (**Fig. 4g**). RANKL signaling has been extensively studied in GCTB (25). The expression of RANKL, a master regulator of osteoclast differentiation (26), has been shown to be upregulated in GCTB stromal cells causing an osteolytic phenotype (25). We verified the increased expression of *RANKL* in H3.3 MUT cells (**Fig. 4h, Fig. S4b**) and additionally observed decreased expression and secretion of its decoy receptor Osteoprotegerin *(OPG, TNFRSF11B)* (**Fig. 4i, Fig. S4 b, c**). Similar expression patterns were already described by us earlier (24). *OPG* had decreased levels of the active histone marks H3K4me3 and H3K27ac potentially indicating a missing activation by a transcription factor (**Fig. S4d**). One known transcription factor of *OPG* is Early B-cell Factor 2 *(EBF2)* (27,28) which is a bivalent gene (**Fig. S4l**) and belongs to key regulators of osteogenic differentiation in mice (29). *OPG* became downregulated after siRNA mediated EBF2 knockdown, confirming a role of EBF2 in *OPG* expression in stromal cells (Fig. 4j, Fig. S4e). The EBF family is a conserved group of four transcription factors. Our RNA-seq analysis found *EBF2* and *EBF3* to be differentially expressed between H3.3 WT and H3.3 MUT stromal cells (Fig. 4k, Fig. S4f). *EBF3*, previously reported as a tumor suppressor in glioblastoma (30,31), showed reduced expression in H3.3 MUT cells whereas EBF2 expression was almost completely lost. We found the EBF2 locus to be hypomethylated with a focal hypermethylation around the promoter region (**Fig. 4l**). Increased H3K9me3 and H3K27me3 levels and decreased levels of H3K27ac supported a repressed state of *EBF2*. Lost ATAC signals in H3.3 MUT cells indicated differentially closed chromatin. These findings link the H3.3 G34W-associated epigenetic dysregulation of *EBF2* expression to the osteolytic phenotype of GCTB.

**Figure 4:**
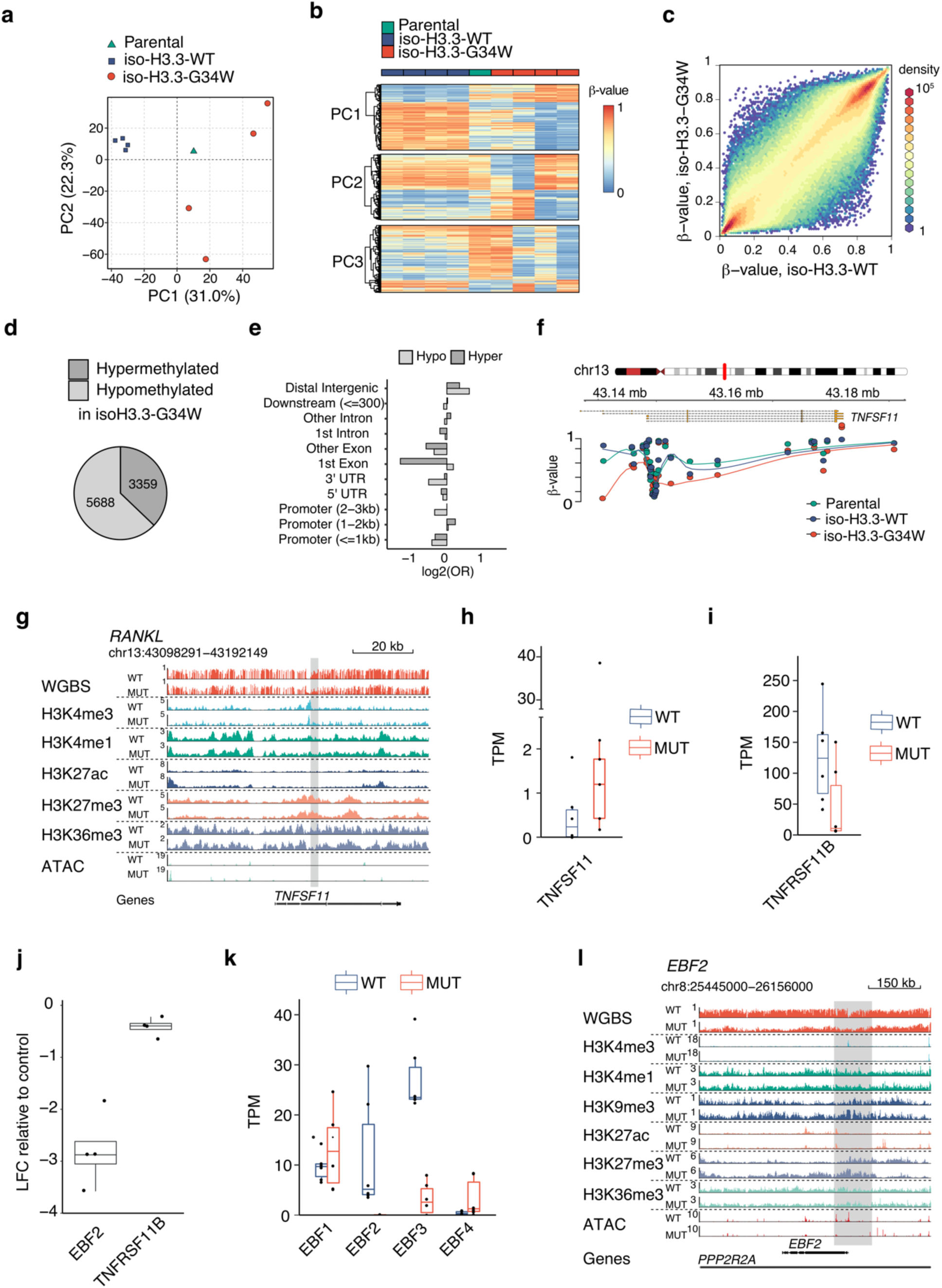
DNA methylation differences can be recapitulated in isogenic Hela cells and link epigenetic distortion to the osteolytic phenotype of GCTB. (on the next page). **a**. Principal component analysis of HumanMethylationEPIC profiles of iso-H3.3-WT and iso- H3.3-G34W HeLa cells. **b**. Methylation levels of top 2000 probes associated with each of the first three principal components in iso-H3.3-WT and iso-H3.3-G34W cells. **c**. Scatter plot of individual CpG probe methylation in iso-H3.3-WT (x-axis) vs. iso-H3.3-G34W (y-axis). Color dots indicate incremental delta means of methylation, with orange representing >0.5 delta-mean. **d**. Fractions of HumanMethylationEPIC probes, differentially methylated between iso- H3.3-G34W and iso-H3.3-WT HeLa cells, by direction of DNA methylation change. **e**. Enrichment (positive values) or depletion (negative values) of major gene model features among differentially methylated probes. OR, odds ratio. **f**. Genome browser snapshot displaying DNA methylation levels of CpGs in the vicinity of the *TNFSF11* locus in iso-H3.3- WT and iso-H3.3-G4W cells. Points represent methylation values individual CpGs and the lines depict LOESS curves with degree 1 and span 0.5. **g**. Genomic browser view of gene *TNFSF11* encoding for RANKL. Each lane (dash-separated) represents normalized averaged signals of DNA methylation, the level of H3K4me3, H3K4me, H3K9me3, H3K27ac, H3K27me3, H3K36me3 and ATAC in several replicates of H3.3 WT (lane-wise top) or H3.3 MUT (lane-wise bottom) cells. **h**. Expression levels of RANKL *(TNFSF11)* in H3.3 WT (blue) H3.3 MUT (red) cells as analyzed by RNA-seq. TPM, transcripts per million. **i**. Expression levels of OPG *(TNFRSF11B)* in H3.3 WT (blue) H3.3 MUT (red) cells as analyzed by RNA-seq. TPM, transcripts per million. **j**. Expression analysis of *EBF2* and OPG *(TNFRSF11B)* by qPCR 48 hours after siRNA-mediated knockdown of EBF2 in H3.3 WT cells (UPI-13). Log fold change (LFC) relative to expression in cells transfected with a control siRNA. Each dot represents one replicate. Boxplot bar represents mean, box gives the IQR, whiskers span additional 1.5*IQR below and above the bars. Results showed significance in a one-sample t-test with p-values < 0.05. **k**. Expression levels of all members of the EBF family in H3.3 WT (blue) H3.3 MUT (red) cells as analyzed by RNA-seq. TPM, transcripts per million. **l**. Genomic browser view of the *EBF2* locus. Each lane (dash-separated) represents normalized averaged signals of DNA methylation, the level of H3K4me3, H3K4me, H3K9me3, H3K27ac, H3K27me3, H3K36me3 and ATAC in several replicates of H3.3 WT (lane-wise top) or H3.3 MUT (lane-wise bottom) cells.

Taken together, we show that the stable introduction of H3.3-G34W into a GCTB unrelated cell line recapitulates the DNA hypomethylation trend seen in GCTB stromal cells. Furthermore, we could detect epigenetic alterations that directly and indirectly affected the expression of key regulators of bone metabolism shedding novel light on the emergence of the osteolytic phenotype in GCTB.

### H3.3 MUT cells exhibits localized changes at bivalent promoters and have a distinct osteogenic differentiation state

Besides dominant hypomethylation in heterochromatic regions, we additionally observed profound epigenetic changes at bivalent domains (**Fig. 2d**), which are well known sites of H3.3 deposition (5,32). Changes included gain of DNA methylation which coincide with decreased accessibility, decrease of H3K27me3 and increased levels of H3K36me3 (**Fig. 2d**). Most promoter-associated ATAC peaks that were lost in H3.3 MUT cells overlapped with bivalent promoters (**Fig. S5a**). As a consequence of epigenetic disturbance, bivalent genes comprised a significant portion of differentially expressed genes both up- and downregulated in H3.3 MUT cells (**Fig. S5b**). It was previously reported that comparable epigenetic and transcriptional perturbations at bivalent genes occur in H3.3-deficient mouse ESCs (7). Genes with decreased expression in H3.3 MUT were significant enriched in Polycomb-target genes, including several developmental transcription factors (**Fig. S5c, Table S4**). This was also supported by genomic overlap enrichment analysis of lost ATAC peaks showing strong enrichment at regions where binding of PRC2 components EZH2 and SUZ12 was found in numerous cell lines (**Fig. S5d, Table S3**). GO analysis of closed regions revealed many categories related to differentiation (**Fig. S5e**). Impaired differentiation was already suggested for tumor entities harboring mutations in histones including GCTB (15,33). To investigate whether H3.3 WT and H3.3 MUT cells differ in their osteogenic differentiation state, we performed a comparison across all genes covered by RNA-seq with those obtained from an *in vitro* differentiation of MSCs. The expression changes during osteogenic differentiation largely correlated inversely with the expression changes observed between H3.3 WT and H3.3 MUT cells, with anticorrelation increasing from very early to late differentiation stages (**Fig. 5a**). Accordingly, an overlap of the differentially expressed genes from both experiments showed that the majority of matches were upregulated during differentiation and downregulated in H3.3 MUT cells (**Fig. S5f**). Examples include insulin growth factor 2 *(IGF2)* and leptin *(LEP)* previously implicated in osteogenic differentiation (34,35) (**Fig. 5a**). A global principal component analysis confirmed a less differentiated state of H3.3 MUT cells resembling MSC in a very early stage of differentiation and H3.3 WT cells showing an expression profile more similar to MSC in early to middle stage of osteogenic differentiation (**Fig. 5b**). To confirm differences in the osteogenic differentiation state, we stained H3.3 WT and H3.3 MUT stromal cells for the activity of the osteogenic marker alkaline phosphatase (ALP). Most of the H3.3 WT cells exhibited ALP activity whereas only some H3.3 MUT cells showed ALP activity (**Fig. 5c**). Quantification of ALP activity relative to viability as a surrogate for cell count confirmed reduced ALP activity for H3.3 MUT stromal cells (**Fig. 5d**). Impaired differentiation of GCTB stromal cells was suggested earlier based on the analysis of histological markers (36) and transcriptomic profiling (37). We performed an osteogenic differentiation of H3.3 MUT and H3.3 WT cells from several patients *in vitro* to analyze the potential of H3.3 MUT and H3.3 WT cells to differentiate. While both H3.3 WT and H3.3 MUT stromal cells showed an increase of ALP activity indicating the potential for osteogenic differentiation (**Fig. 5c** and **d**), we noticed that H3.3 MUT stromal cells lagged behind and did not achieve the level of ALP activity reached by H3.3 WT stromal cells (**Fig. 5c** and **d**). We conclude that H3.3-G34W associates with changes in bivalent regions and that their deregulation potentially has an effect on osteogenic differentiation. H3.3 WT and H3.3 MUT stromal cells therefore differ in their state of osteogenic differentiation in GCTB patients and this difference contributes to the herein found epigenetic alterations (**Fig. 5e**).

**Figure 5:**
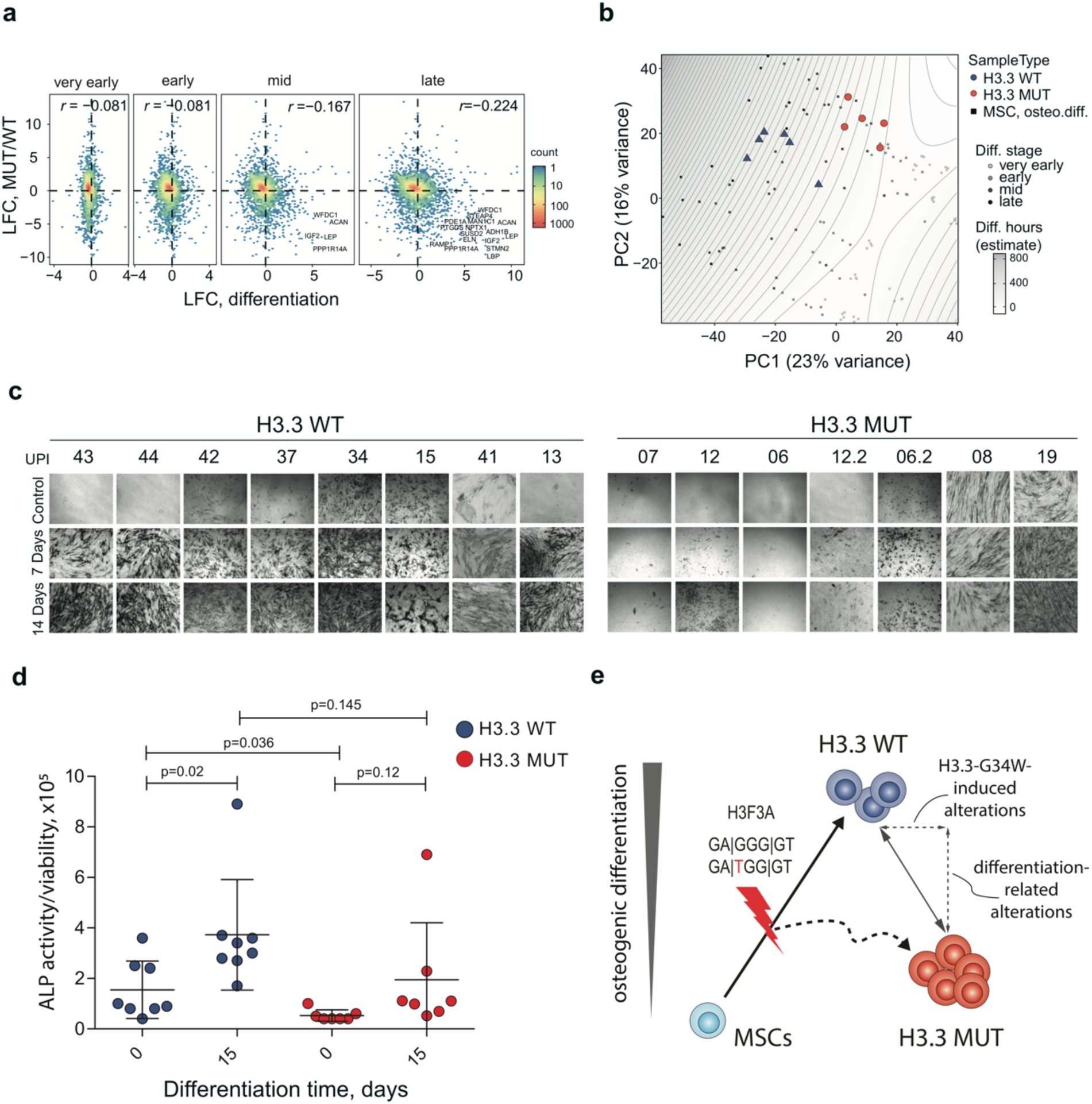
Impairment of osteogenic differentiation in H3.3 MUT stromal cells. **a**. Correlation between differentiation- and H3.3 G34W-associated gene expression changes. For the differentiation log-fold changes (LFC) were calculated between average of profiles from each differentiation timepoint group (very early: 0.5 to 2 hours; early: 4 to 554 16 hours, mid: 24 to 168 hours; and at: 336 to 504 hours) and the profiles of MSCs sampled prior to the start of the experiment. Each hexagon reflects density of the underlying data points. Genes with most pronounced changes in both data sets are marked. **b**. Principal component analysis of RNA-Seq data from osteogenic differentiation of MSC and expression profiles of GCTB samples added via a projection onto the subspace spanned by principal components 1 and 2. The sampled differentiation timepoints were grouped as: very early (0.5 to 2 hours), early (4 to 16 hours), mid (24 to 168 hours) and late (336 to 504 hours). **c**. Alkaline phosphatase staining of nt-SC, H3.3 WT and H3.3 MUT cells during osteogenic differentiation. **d**. Quantification of alkaline phosphatase activity relative to the viability as a surrogate for cell count over the time course of 14 days of *in vitro* osteogenic differentiation. **e**. Model for GCTB tumorigenesis: The *H3F3A* mutation encoding H3.3-G34W occurs in osteoblastic precursor cells and leads to alterations in osteogenic differentiation. Epigenetic differences described between H3.3 WT and H3.3 MUT stromal cells are the result of measuring cells at different differentiation stages and direct effects of the mutated histone.

## Discussion

Mutant epigenetic regulator proteins, including histones, have been described in multiple cancer types (2). Despite the fact that oncogenic mutations in histones influence the epigenetic landscape, deep insight into the mechanistic ramifications to cancer initiation is still incomplete. Several studies investigated the mechanism of how the G34W substitution in histone variant H3.3 affects the epigenetic machinery responsible for its post-translational modifications. To this end, reduced H3K36 methylation and increased H3K27me3 *in cis* was described in model systems and verified by us in this work (16,38). However, whether these effects can be found in patients and whether epigenetic changes are involved in tumorigenesis of GCTB was not known so far. In this paper, we therefore analyzed GCTB tissue as well as patient-derived primary stromal cells from four different centers to investigate H3.3-G34W-associated epigenetic changes and their contribution to the GCTB neoplasia. The high incidence of H3.3-G34W in GCTB in a largely unaltered genomic context leaves GCTB as a suitable system to study histone mutation-driven tumorigenesis.

We found that, despite that H3.3-G34W is incorporated into chromatin, its presence did not lead to changes in the global amount of H3K36me2/me3 as shown for other substitutions affecting H3.3 lysine 27 and lysine 36 (11,13,39). This ruled out a possible *in trans* effects of H3.3-G34W on histone posttranslational modifications analyzed in this study. Using ChIP sequencing, we identified high confidence H3.3-G34W enrichment sites in patient cell lines and analyzed common histone marks at these regions. No pronounced differences were found that could recapitulate the *in cis* effects on H3.3 lysine methylation observed in HEK293 cells. The absence of clear-cut *in cis* effects upon chromatin modifications could be explained by an inability to distinguish wild-type and mutant H3.3 in GCTB samples by western blot analysis. In ChIP analysis, nucleosomes of mutant cells could include both, wild-type as well as mutated H3.3 obliterating possible *in cis* effects. Moreover, antibodies against histone modifications cannot distinguish between H3 variants and the overall small fraction of the mutant H3.3 (25% of overall H3.3) in the total H3 pool makes analysis of H3.3 modifications challenging.

We show that H3.3-G34W, the sole recurrent alteration of GCTB, associates with large-scale differences in multiple epigenetic marks. H3.3 MUT stromal cells show heterochromatic defects, specifically reduced genome-wide levels of DNA methylation and gained accessibility at heterochromatic regions which is in line with previous reports on pediatric glioblastomas with the G34R substitution in H3.3 (39). These changes could potentially contribute to the genomic instability described for GCTB (23). We furthermore observed localized changes at bivalent regions, some of the same sites previously described as targets of H3.3 deposition (5,7). DNA methylation changes could be recapitulated using an isogenic system verifying effects on the methylome to be H3.3-G34W-specific. Moreover, we show H3.3-G34W-associated methylome changes to directly or indirectly affect the expression of the main players of bone metabolism, RANKL and OPG. These results connect the H3.3-G34W-associated epigenetic dysregulation to the osteolytic phenotype, a hallmark of GCTB.

While the epigenomic differences between H3.3 WT and H3.3 MUT stromal cells can be partially ascribed to direct effects of H3.3-G34W, other alterations observed can be explained by the fact that H3.3 MUT and H3.3 WT stromal cells represent distinct differentiation stages. Assuming epigenetic reprogramming during differentiation as demonstrated in recent studies (40,41), this could also contribute to the epigenetic differences seen between H3.3 WT and H3.3 MUT cells. While expression profiles from H3.3 MUT cells resemble precursor states of osteoblasts, the H3.3 WT cells resemble more mature osteoblasts, suggesting impaired or delayed differentiation of H3.3 MUT cells as already suggested by (42). A weak differentiation signature is in line with previous reports suggesting that neoplastic GCTB stromal cells are expressing markers of an early osteoblastic differentiation (43). Genes such as insulin growth factor 2 *(IGF2)* (34), leptin *(LEP)* and stathmin-like 2 *(STMN2)*, found to be dysregulated in H3.3 MUT stromal cells, were described to play a role in mesenchymal stem cell differentiation and osteoclastogenesis in mice, in promoting bone differentiation (35) and as a marker for osteogenesis (44), respectively. The impairment of differentiation of H3.3 MUT stromal cells could be mediated by H3.3-G34W-associated downregulation or prevention of induction of genes upregulated during osteogenic differentiation through a yet unknown mechanism. Epigenetic repression of bivalent genes and dysregulation of PRC2 targets described in this study could potentially contribute to the observed phenotype. H3.3 MUT stromal cells are still able to undergo osteogenic differentiation, as it was also found in studies not separating H3.3-mutant neoplastic from H3.3 -wild-type stromal cells of GCTB tissue (45,46).

Taken together, our results suggest that the molecular mechanism behind H3.3-G34W-induced epigenomic alterations and neoplastic transformation in GCTB is different to the action of other mutant histone variants such as H3.3-K27M and H3.3-K36M, where the mutated amino acid is a direct target for histone modification (11,13,15). This suggests that the H3.3-G34W substitution alters the N-terminal interactome as previously described (24), recruiting novel binders or disrupting interactions with binders. The described DNA methylation changes can be indicative of H3.3-G34W affecting either binding or activity of enzymes writing (DNMTs) or erasing (TETs) this modification. On the structural side it is known that DNMT3a/b recognize H3K36me3 with their PWWP-domains (47,48). The G34W substitution might interfere with binding of DNTM3a/b PWWP preventing proper establishment of DNA methylation patterns. The thereby induced remodeling of the epigenome associated with H3.3-G34W, in particular changes at heterochromatic and bivalent regions, could contribute to an impaired osteogenic differentiation and the phenotypes of GCTB: stochastic chromosomal rearrangements and increased RANKL signaling. Precise sequences of molecular events leading to these phenotypes will be a matter of further studies.

## Methods

### Patient samples

Biopsies and derived primary cell lines were obtained from patients in the Orthopedic University Hospital, Heidelberg (UHOK), the Korean Cancer Center (KCC), the University Clinic of Leipzig (UKL) and the University Medical Center Hamburg-Eppendorf (UKHE). The complete list of samples used in the study is available in **Table S1**. The use of patient samples and the experiments performed in this study was approved by and in accordance with guidelines and regulations by the Ethics Committees of the University of Heidelberg, University Clinic of Leipzig, University Medical Center Hamburg-Eppendorf, and the National Cancer Center of Korea (IRB NCC2015-0070). All H3.3 WT and H3.3 MUT cells analyzed in the manuscript were obtained from different patients or indicated as relapse samples by addition of .2 to the UPI.

### Isolation of GCTB stromal cells from patient tissue

Tumor tissue from surgical resections was mechanically cut into small pieces and digested with 1.5 mg/ml collagenase B (Roche Diagnostics, Mannheim, Germany) at 37°C for 3 h in Dulbecco’s Modified Eagle Medium (DMEM) (Lonza GmbH, Wuppertal, Germany) high glucose supplemented with 10% fetal calf serum (FCS) (Biochrom, Berlin, Germany), and 100 U/ml penicillin/streptomycin (Lonza GmbH). Cells were collected by centrifugation, washed twice in PBS and cultured in DMEM as described above. Twenty-four hours after plating, cells were carefully treated with Trypsin/EDTA (Lonza GmbH) leaving the giant cells attached in the culture flask. Detached cells were cultured for further 2 passages to eliminate histiocytes and remaining giant cells.

### Isolation of nt-SC cell lines from the bone marrow

Nt-SCs were isolated from fresh bone marrow samples derived from the iliac crest under the approval of the Ethics Committee of the University of Heidelberg. Bone marrow cells were purified on a Ficoll-Paque™ Plus density gradient (GE Healthcare, München, Germany), washed in PBS and treated with erythrocyte lysis buffer (0.154 M NH_4_Cl, 10 mM KHCO_3_, 0.1 mM EDTA) to remove erythrocytes. The nt-SC-enriched fraction was seeded and cultured in DMEM high-glucose supplemented with 12.5% FCS (Biochrom), 2 mM L-glutamine, 100 units/ml penicillin, 100 μg/ml streptomycin (Lonza), 4 ng/ml basic fibroblast growth factor (Merck Chemicals GmbH, Darmstadt, Germany, 50 μM 2-mercaptoethanol and 1% non-essential amino acids (Invitrogen, Karlsruhe, Germany). After 48 h, cultures were washed with PBS to remove non-adherent material. During expansion, medium was replaced twice a week.

### Cell line maintenance

The GCTB stromal and nt-SC cell lines were cultivated in DMEM/F12 no phenol red (Gibco, Invitrogen, Life Technologies, Paislay, UK) supplemented with 10% FCS (Biochrom, Berlin, Germany) and 2 ng/μl basic FGF (Biolegend, San Diego, USA) at 37°C and 5% CO_2_. To avoid contact inhibition, cells were split after reaching maximum 90% of confluency.

### Isogenic cell lines

The establishment of H3.3-GFP isogenic cell lines in HeLa cells utilized the zinc-finger (ZF) targeting methodology as previously described (24). In brief, after transfection with ZF and targeting constructs with electroporation, cells were cultured in media under neomycin selection using G418 for four weeks. Surviving cells were FACS-sorted and individual clones were allowed to propagate. HEK293T were lentivirally-transduced with human H3.3-HA-3XFLAG constructs in pLVX expression vectors and selected with puromycin. A pLVX-AcGFP construct was used as control. Cells were maintained in DMEM (Invitrogen™ Life Technologies, Carlsbad, USA) supplemented with 10% FCS (Biochrom, Berlin, Germany) and penicillin-streptomycin.

### Immunohistochemistry

For immunohistochemical detection of the H3.3-G34W mutation formalin fixed paraffin embedded GCT tissue sections were deparaffinized in Roti-Histol (Carl Roth GmbH, Karlsruhe, Germany) and rehydrated in isopropanol. Antigen retrieval was performed using Dako target retrieval solution pH 6 (Dako, Hamburg, Germany) for 5 min at 121°C in a pressure cooker. Sections were blocked for 30 min at room temperature using PBS supplemented with 5 % bovine serum albumin (BSA). The primary rabbit anti-H3G34W antibody (Active Motiv, Carlsbad, USA) was diluted 1:1000 in PBS/1% BSA and incubated over night at 4°C. The signal was amplified using the BrightVision +Poly-AP kit (VWR, Darmstadt, Germany) according to the manufacturer’s instructions. Samples were counterstained with hematoxylin (Carl Roth GmbH) and mounted using Neo-Mount (Merck, Darmstadt, Germany). All used antibodies are listed in **Table S5**.

### Lineage verification by flow-cytometric analysis

Stromal cells were harvested by trypsination and transferred to 1,5 ml Eppendorf in PBS. Cells were washed with PBS 2% FCS and centrifuged for 5 min at 250 g at 4°C. The antibody staining cocktail was prepared in PBS 2% FCS and cells were stained 20 min at 4°C (all antibodies are specified in the **Table S5**). Afterwards cells were washed with PBS 2% FCS to remove unbound antibodies and centrifuged at 250 g for 5 min. To exclude dead cells during flow cytometric analysis, propidium iodide was added prior to flow analysis. Data were acquired on a BD FACSAria Fusion (Beckton, Dickinson & Company (BD)) and data were analyzed using FlowJo (BD). Prism (GraphPad Software) was used to generate bar graphs. Hematopoietic/erythroid cells were defined as CD45+ CD235+, CD45-CD235-CD105+ and CD45-CD235-CD90+ cells were defined as mesenchymal cells. Freshly frozen iliac crest bone marrow aspirates from healthy donors were used as healthy controls.

### OPG expression quantification with enzyme-linked immunosorbent assay (ELISA)

5000 cells were seeded into a 96-well plate and cultivated in 100μl media for two days. ELISA was performed with the Abcam human Osteoprotegerin ELISA Kit following the manufacturer’s instructions.

### Cell titer blue assay

Cells were cultivated in a 96-well plate. 100μl fresh media (DMEM/F12 no phenol red (Gibco, Invitrogen, Life Technologies, Paislay, UK) supplemented with 10% FCS (Biochrom, Berlin, Germany)) were added and supplemented with 20μl CTB reagent from Promega. After 2.5h of cultivation at 37°C Fluorescence was recorded at 560Ex/590Em.

### siRNA knockdown

120.00 cells were seeded into a 6-well plate. After 24 hours, medium was changes and transfection was performed using 1μl Dharmafect1 (Horizon Discovery, <u>Waterbeach, UK</u>) reagent in combination with 2 μl of a 25 μM siRNA SMARTpool of Dharmacon (Horizon Discovery, <u>Waterbeach, UK</u>) targeting EBF2. Cells were harvested 48 hours after transfection.

### Detection of H3F3A-G34W mutation by mutation-specific PCR

Genomic DNA was isolated using the Quick DNA Miniprep kit (Zymo research, Freiburg, Germany) according to the manufacturer’s protocol. PCR amplification was performed using H3F3A wild-type and H3F3A-G34W specific primer, respectively. The reaction consisted of 2 U Platinum Taq polymerase (Thermo Fisher Scientific, Dreieich, Germany), 0.6 μl MgCl_2_ (50 mM), 0.4 μl dNTPs (10 mM each), 0.5 μl of each primer (10 μM) and 100 ng genomic DNA as template in a total volume of 20 μl. Samples were incubated at 94°C for 3 min followed by 34 cycles of denaturation at 94°C for 15 s, annealing at 66°C for 20 s and extension at 72°C for 30 s and a final extension step at 72°C for 7 min. PCR products were separated on a 1.6% agarose gel, visualized by Midori Green (Biozym, Hessisch Oldendorf, Germany) and imaged. All primers are listed in **Table S6**.

### Detection of H3F3A-G34W mutation by Sanger sequencing

DNA was extracted using the Quiamp Mini Kit (Qiagen, Hilden, Germany), according to the manufacturer’s instructions. The PCR reaction to amplify the mutation spanning *H3F3A* region consisted of 1U Hot Star Taq polymerase (Thermo Fisher Scientific, Dreieich, Germany), 0.8 μl dNTPs (10 mM each, Fermentas, St. Leon-Rot, Germany), 2 μl of each primer (10 μM, Sigma-Aldrich, Taufkirchen, Germany) and 100 ng genomic DNA as template in a total volume of 40 μl. Samples were incubated at 95°C for 15 min followed by 10 cycles of denaturation at 94°C for 45 s, annealing at 61-56°C for 30 s (touchdown -0.5°C/cycle) and extension at 72°C for 60 s followed by 25 cycles of denaturation at 94°C for 45s, annealing at 56°C for 30 s and extension at 72°C for 60 s and a final extension step at 72°C for 10 min. Sequencing was performed by GATC Biotech AG, Konstanz, Germany. All primers are listed in **Table S6**.

### Deep targeted resequencing using MiSeq

Deep resequencing of *H3F3A* amplicons was performed as earlier described (49). In brief, DNA was extracted using the Qiamp Mini Kit (Qiagen, Hilden, Germany), according to the manufacturer’s instructions. The PCR reaction to amplify the mutation spanning region of the *H3F3A* gene consisted of 0.35U HotStart Q5 polymerase (NEB, Ipswich, USA), 0.6 μl dNTPs (10 mM each, Fermentas, St. Leon-Rot, Germany), 0.3 μl of each primer (10 μM, Sigma-Aldrich, Taufkirchen, Germany) and 25 ng genomic DNA as template in a total volume of 25 μl. Samples were incubated at 98°C for 1 min followed by 33 cycles of denaturation at 98°C for 10 s, annealing at 61°C for 30 s and extension at 72°C for 20 s, and a final extension step at 72°C for 2 min. Primer sequences are listed in **Table S6**. Primers included a sequence complementary to the primers used for library preparation. Samples were separated on a 1.2% agarose gel and DNA was visualized by Ethidium Bromide. Gel extraction was performed using the Gel extraction Kit (Qiagen, Hilden, Germany). Libraries were prepared using 12.5μl NEB Next HF 2x PCR mix (New England Biolabs, USA) in combination with 0.75 μl of 10 μM IDT primers with Nextera handles and TruSeq Unique-Dual Indices (IDT, USA), 0.3 μl 100x Sybr green and 11 μl of DNA (2ng). Amplification was performed for 6 cycles with the following progression: 98°c 30sec, 98°C 10 sec, 62°C 30 sec and 72°C 15 sec. Libraries were pooled and sequenced on a single flow-cell lane in a or 300 bp paired-end MiSeq run (Illumina, San Diego, CA, USA). The reads were demultiplexed using custom scripts, aligned to the GRCh37 assembly using *bwa mem* (v. 0.7.8) (50) with default settings. Base frequencies were read into R and analyzed using package *deepSNV* (v. 1.24.0) (51).

### Detection of genomic structural variations using multiplex fluorescence in situ hybridization, M-FISH

M-FISH was performed as described in (52). Briefly, seven pools of flow-sorted whole chromosome painting probes were amplified and directly labelled using DEAC-, FITC-, Cy3, TexasRed, and Cy5-conjugated nucleotides or biotin-dUTP and digoxigenin-dUTP, respectively, by degenerative oligonucleotide primed (DOP)-PCR. Prior hybridization, metaphase preparations of the H3.3 WT and H3.3 MUT cells were digested with pepsin (0.5 mg/ml; Sigma) in 0.2N HCL (Roth) for 10 min at 37°C, washed in PBS, post-fixed in 1% formaldehyde, dehydrated with a degraded ethanol series and air dried. Slides were denatured in 70% formamide/1 x SSC/15% dextran sulfate for 2 min at 72°C. Hybridization mixture consisting of 50% formamide, 2 x SSC, Cot1-DNA, and labeled DNA probes was denatured for 7 min at 75°C, preannealed for 20 min at 37°C, and hybridized to the denatured metaphase preparations. After 48 hours incubation at 37°C slides were washed at room temperature in 2 x SSC, 3x 5 min, followed by 2x 5 min in 0.2% SSC/0.2% Tween-20 at 56°C. For indirect labeled probes, a two-step immunofluorescence detection was carried out using biotinylated goat anti avidin followed by streptavidin Laser Pro IR790, and rabbit anti digoxin followed by goat anti rabbit Cy5.5. Slides were washed in 4 x SSC/0.2% Tween-20, counterstained with 4.6-diamidino-2-phenylindole (DAPI) and covered with antifade solution. Images of 20 metaphase spreads of the H3.3 WT and H3.3 G34W cells were captured for each fluorochrome using highly specific filter sets (Chroma technology, Brattleboro, VT), and processed using the Leica MCK software (Leica Microsystems Imaging Solutions, Cambridge, UK), respectively.

### Chromatin fractionation

Cell pellets were resuspended in lysis buffer (10 mM HEPES pH 7.6, 10 mM KCl, 0.05% NP40) with protease inhibitors and incubated on ice for 30 minutes. Samples were centrifuged at 13200 rpm at 4°C for 10 min and the supernatant was taken as cytosolic fraction. Leftover pellet was further lysed with low salt buffer (10 mM Tris HCl pH 7.5, 3 mM MgCl_2_) supplemented with 1% Triton X-100 for 15 min on ice and centrifuged at 13200 rpm at 4°C for 10 min. Supernatant was taken as nuclear proteins. Leftover pellet was resuspended in 0.2M HCl, incubated on ice for 20 min and centrifuged at 13200 rpm at 4°C for 10 min. The supernatant was neutralized with 1M Tris HCl buffer (pH8) and used as the chromatin fraction.

### Western blot

Proteins were separated using SDS polyacrylamide gel electrophoresis (SDS-PAGE) and analyzed by Western blotting using the antibodies listed in the **Table S5**. Chemiluminescence signals were imaged using Amersham Imager 680 (GE, Boston, USA). For the separation of endogenous histone H3 proteins and ectopically-expressed H3.3-HA-3XFLAG in HEK293T, we used 8-16% Mini-PROTEAN® TGX gels (Bio-Rad Laboratories).

### Differentiation and alkaline phosphatase staining

50.000 cells were seeded in 24-well plates. When confluent medium was changed to osteogenic differentiation medium: DMEM high Glucose mit L-Glutamin supplemented with 10%FCS, 0.1 μM Dexamethason, 0.17mM Ascorbinsäure-2-phosphat and 10mM ß-Glycerophosphat. Alkaline phosphatase activity was analyzed using BCIP/NBT Alkaline Phosphatase Substrate Kit (Vector Laboratories, Burlingame, CA, USA) after 1min of 4% PFA fixation.

### Quantification of ALP activity

50.000 cells were seeded in 24-well plates. When confluent media was changed to differentiation media (compare Differentiation and alkaline phosphatase staining) and the first measurement was performed. After CTB as described above, wells were PBS washed and fixated for 60sec with 4% PFA in PBS. After PBS washing 500 μl Alkaline Phosphatase Yellow (pNpp) Liquid Substrate (System for ELISA, Sigma-Aldrich, USA) was added and incubated at 37°C for 8 minutes. Absorbance was measured at 405nm.

### Whole-genome sequencing and analysis

Read pairs were mapped to the human reference genome (build 37, version hs37d5), using *bwa mem* (v. 0.7.8) (50) with minimum base quality threshold set to zero [-T 0] and remaining settings left at default values, followed by coordinate-sorting with *bamsort* (with compression option set to fast REF1) and marking duplicate read pairs with *bammarkduplicates* (with compression option set to best (53)); both are part of *biobambam* package (v.0.0.148) (54). Somatic SNVs were identified with the DKFZ SNV-calling workflow (55). Somatic SNVs and indels in matched tumor normal pairs were identified using the DKFZ core variant calling workflows of the ICGC Pan-cancer Analysis of Whole Genomes (PCAWG) project (https://dockstore.org/containers/quay.io/pancancer/pcawg-dkfz-workflow). Tumor and matched control samples were analyzed by *Platypus* (v. 1.0) (56) to identify indel events. SNVs and indels from all samples were annotated using *ANNOVAR* (v. 2017Jul16) (53) according to GENCODE gene annotation (v. 19) and overlapped with variants from dbSNP10 (build 141) and the 1000 Genomes Project database. Genomic structural rearrangements were detected using *SOPHIA* (v.34.0) (https://bitbucket.org/utoprak/sophia/src) as described in (57). Briefly, *SOPHIA* uses supplementary alignments as produced by *bwa mem* as indicators of a possible underlying SV. SV candidates are filtered by comparing them to a background control set of sequencing data obtained using normal blood samples from a background population database of 3261 patients from published TCGA and ICGC studies and both published and unpublished DKFZ studies, sequenced using Illumina HiSeq 2000 (100 bp), 2500 (100 bp) and HiSeq X (151 bp) platforms and aligned uniformly using the same workflow as in this study. An SV candidate is discarded if (i) it has more than 85% of read support from low quality reads; (ii) the second breakpoint of the SV was unmappable in the sample and the first breakpoint was detected in 10 or more background control samples; (iii) an SV with two identified breakpoints had one breakpoint present in at least 98 control samples (3% of the control samples); or (iv) both breakpoints have less than 5% read support. Statistics over SVs for 9 samples with matched control and integrated variant analysis over all samples were based on *SOPHIA* calls. Allele-specific copy-number aberrations were detected using *ACEseq* (v. 1.0) (58). SVs called by *SOPHIA* were incorporated to improve genome segmentation.

### ATAC-sequencing and analysis

Libraries for ATAC-sequencing were prepared as previously published with modifications (59). Briefly, cells were lysed by 1% NP40 and tagmented at 55°C for 8 minutes in a reaction mix with 2.5μl of TDE1 (Nextera Illumina DNAKit), 25μl Tagmentation buffer (Nextera Illumina DNA Kit) and 25μl of lysed cells. Reaction was stopped by adding 10 μl Guanidium (5 M) and samples were purified using Ampure Beads. Libraries were generated using NEBNext High Fidelity PCR Mix and sequenced on the Illumina HiSeq 2000 platform. Sequencing reads were adaptor-trimmed using *cutadapt* (v. 1.10) (60). Genomic alignments were performed against the human reference genome (hg19, NCBI build 37.1) using *Bowtie2* (v. 2.3.0) (61). The non-default parameters “-*q 20 -s”* were used. PCR duplicates were removed by *PicardMarkDuplicates* (v. 1.125). Signal tracks were generated using *deepTools* (v. 2.3.3) (62). Peaks were called using *Macs2* (v. 2.1.1.) (63) with the parameters “--*nomodel --shift -50 --extsize 100 --qvalue 0.01”*. All peaks were merged to create a common bed file with read counts before differential analysis using *edgeR* (v. 0.3.16) (64). Gene annotations were made using *ChIPpeakAnno* (v. 3.18.0) (65). Transcription binding motif analysis was performed using *HOMER* (v. 4.9) (66). Motifs with a *P*-value < 0.01 and a ratio of motif to background above 1.1 were defined as significantly enriched.

### ChIP-sequencing and analysis

We used the ChIP-mentation protocol (67) to map the genomic distribution of WT and G34W H3.3, total H3, as well as 6 histone modifications (H3K4me1, H3K4me3, H3K9me3, H3K27ac, H3K27me3 and H3K36me3) in a subset of samples (see details in **Table S1**). To validate the specificity of the H3.3 and H3.3 G34W antibody used for ChIP-Seq analysis, we performed a validation experiment with a Histone code peptide array (JPT, Berlin, Germany) containing short peptides that densely cover most of the known histones and their modifications. We did not observe any significant binding of the G34W-speicifc antibody to H3.3 peptides and a highly specific binding of the H3.3 antibody. Sequence reads were preprocessed using *cutadapt* (v. 1.10) (60) and aligned with *Bowtie2* (v. 2.3.0) (61) with the default command line options. We used *deepTools* (v. 2.3.3) (62) with non-default options *“--binSize 10 --extendReads 400 – -normalizeTo1x 2451960000 --ignoreForNormalization chrX”* to quantify genomic coverage in fixed-size window intervals for meta-plots and heatmaps.

### Calling of H3.3-G34W enriched regions

We used the Poisson test-based binarization module of the ChromHMM software (v. 1.18) (68) to generate 200bp windows with statistically significant enrichment of the wild-type H3.3 or H3.3-G34W signal over a simulated background. Adjacent windows were merged to generate primary enrichment regions. The regions were filtered against a union of the ENCODE ChIP-seq blacklists.

### Whole-genome bisulfite sequencing and analysis

DNA sequencing libraries were prepared using the TruSeq NanoDNALibrary PrepKit (Illumina, San Diego, CA, USA) following the manufacturer’s instructions. Paired-end sequencing (2×150 bp) was performed using one lane of a HiSeqX (Illumina) for every sample. Basic statistics about the sequencing results is given in **Table S1**. Raw reads were processed using *Trimmomatic* (v. 0.36) (69) and aligned against reference sequence of the Genome Research Consortium (v. 37) using *bwa mem* (v. 0.7.8) (50) with default parameters, except for invoking *“-T0”*. After alignment duplicates were marked by applying *Picard MarkDuplicates* (v. 1.125). Methylation calling was performed with *MethylDackel* (v. 0.3.0). Coverage filtering still missing). *BSmooth* was used (v. 1.4.0) with default parameters to smooth the methylation profiles in all samples (70). We then used we *DSS* (v. 2.27.0) (71) to call DMRs for pairwise comparison between H3.3 WT and H3.3 MUT cells. Regions with at least 3 CpGs, a minimum length of 50bp and a Benjamin-Hochberg corrected *P* value < 0.05 were selected. All DMRs were filtered requiring a minimal mean methylation-value difference of 0.1.

### HumanMethylation450 and HumanMethylationEPIC analysis

Methylation analysis using HumanMethylation450 arrays was performed by the Genomics and Proteomics Core Facility according to the manufacturer’s instructions. Profiling of isogenic HeLa cell lines with HumanMethylationEPIC arrays was conducted at Korean Cancer Center according to the manufacturer’s instructions. Unnormalized signals (IDAT files) were loaded into R using RnBeads software (v. 2.2.0) (72) and subjected to preprocessing with default option settings. 10,000 sites most variable across all samples were used for both, Principal Component Analysis and clustering analysis, visualized as a heatmap.

### RNA-sequencing and analysis

Poly-A RNA sequencing of GCTB samples was performed according to the standard protocol published elsewhere (73). In brief, total RNA was prepared for each cell line by using the RNAeasy Mini kit (Qiagen, Hilden, Germany) and library preparation was done using TruSeq Stranded mRNA Kit (Illumina), according the manufacturer’s instruction. Paired-end 125bp sequencing runs were performed on Illumina Hiseq2000 v4 machines. Raw sequence reads were preprocessed using *cutadapt* (v. 1.10) (60) to remove sequencing primers and adapters. Reads were aligned to the GRCh37 human reference genome with *HISAT2* (v. 2.0.4) (74) with additional non-default parameters *“--max-intronlen 20000 –no-unal --dta”*. Transcripts were assembled and quantified with *StringTie* (v. 1.3.3) (75) with the GRCh37 transcript database. Differential expression analysis was performed using *DeSeq2* (v. 1.18.1) (76). Genes were called differentially expressed at FDR 0.05 and the absolute log-fold difference of greater or equal to one. To compare expression with the data from all TCGA cohorts, the raw data was alternatively quantified with *kallisto* (v. 0.43.1) (77), and log2-transformed counts were combined with identically processed data downloaded from UCSC Xena portal.

### RNA-Seq of the differentiation samples

RNA-seq of the MSC differentiation samples was performed according to the following protocol. 50,000 cells were seeded in 6-well plates in maintenance medium: MesenPRO-RS™ (Thermo Fisher Scientific, Massachusetts, USA). After 3 days, medium was replaced with osteogenic differentiation medium: DMEM high glucose supplemented with 10% FBS, 1x non-essential amino acids (NEAA), 2mM L-glutamine, 0.28mM ascorbic acid, 10 mM β glycerophosphate, and 10 nM dexamethasone (Sigma-Aldrich, USA). At each time-point: 0, 0.5, 1, 2, 4, 6, 8, 12, 16, 24, 48, 72, 125, 168, 336, 504 (hrs) during osteogenesis, total RNA was isolated using TRIzol (Invitrogen™ Life Technologies, Carlsbad, USA) with the Directzol RNA kit (ZymoResearch, USA) according to the manufacturer’s instruction. RNA-seq library preparation was carried out using the NEBNext Poly(A) mRNA Magnetic Isolation Module and NEBNext Ultra Directional RNA Library Prep Kit for Illumina (New England Biolabs, USA) according to the manufacturer’s instruction. The quantity and quality of the cDNA library were assessed using the Agilent 2200 tapestation (Agilent Technologies, Santa Clara, USA). Paired-end sequencing of the pooled library was carried out using the Illumina NextSeq 500 v2 kit (Illumina, San Diego, USA) according to the manufacturer’s instruction.

### qRT-PCR expression analysis

Total RNA was extracted using the RNeasy Mini kit (Qiagen, Hilden, Germany). cDNA was synthesized using random hexamers (Qiagen, Hilden, Germany), and Superscript III Reverse Transcriptase (Invitrogen, Life Technologies, Paislay, UK) according to the manufacturer’s instructions. qPCR mixture consisted of 3.5 μl Light Cycler 480 Probe master (Roche Diagnostics, Mannheim, Germany) 1 μl 10 μM Primer mix (Sigma-Aldrich, Taufkirchen, Germany) and 0.05μl UPL probe (Roche Diagnostics, Mannheim, Germany). 2.5 μl of a 1:10 dilution of cDNA served as template. Expression analysis was performed on the LightCycler 480-2 (Roche) system with the following progression: 10 min 95°C and 45 cycles of 10 s 95°C, 20 s 55°C, 1 s 72°C. Alternative to the UPL system, Sybr green qPCR was performed in a total volume of 10 μl including 5μl Prima Quant Mix (Steinbrenner, Wiesenbach, Germany), 0.6 μl Primermix (10 μM each, Sigma-Aldrich) and 2 μl of 1:10 dilution of the template cDNA. Samples were incubated at 95°C for 15 min and 15 s at 95°C, 30 s at 55°C and 10 s at 72°C for 45 cycles. Target gene expression was normalized to the housekeeping gene GAPDH using the DCT method (relative expression is equal to 2^-ΔCT^). All used primers are listed in **Table S6**.

### Targeted DNA methylation analysis using MassARRAY

Bisulfite treatment was performed with the EZ DNA Methylation Kit from Zymo Research following the manufacturer’s protocol. PCR was performed with primers listed in **Table S6**using the Qiagen HotStar Taq (Qiagen, Hilden, Germany). The Shrimp alkaline phosphatase step and in vitro transcription were performed with the EpiTYPER Reagent Set from Agena Bioscience following the manufacturer’s protocol. Analyses were performed on the Sequenom Platform (Agena Bioscience, San Diego, USA).

### Gene and genomic feature annotations

Unless specified otherwise, Ensembl transcript and gene annotations were used for the GRCh37 assembly (build 87). List of bivalent genes in H3.3 WT was compiled by overlapping the consensus H3.3 WT peaks of H3K27me3 and H3K4me3 within 2kb from a RefSeq TSS. A consensus list of bivalent genes in human ESCs was obtained from (78). A list of PRC2 target genes was found in (79). A comprehensive list of imprinted genes was obtained from (80). Replication timing domains for the annotation of LMDs originates from Repli-Seq data of (81), processed and publicly deposited by *RepliScan* package (82). MSC-specific chromatin states were taken from the 15-state *ChromHMM* model (68) for bone-marrow derived MSCs generated by the Roadmap Epigenomics consortium (21) (sample E026).

### Identification of large-scale methylation domains

DNA methylation data was summarized in 20 kb tiling windows to eliminate the small-scale variability (e.g. related to CpG islands). The changepoints were then called using R package *changepoint* (v. 2.2.2). DNA methylation data was summarized in the obtained segments and the latter were clustered using standard hierarchical clustering resulting in 6 stable clusters. After clustering adjacent segments that belonged to the same cluster were merged. Two smallest clusters were removed since one contained less then 10 segments and the other one exclusively Y-chromosome segments.

### Genomic overlap enrichment analysis

We tested the significance of overlap of differential ATAC-seq peaks, H3.3 G34W incorporation regions and other regions of interest with publicly available genomic annotations using *LOLA* (v. 1.8.0) (83). In the case of ATAC peaks a union of all called peaks in H3.3 WT and H3.3 MUT was used as background to test enrichments at H3.3 G34W gained and lost peaks relative to each other. For H3.3 WT and G34W enrichment regions the background set consisted of regions called for wild-type H3.3 in H3.3 WT and H3.3 MUT groups, as well as the H3.3-G34W regions. We used the *LOLA* core database for most of the analyses. Enrichment of repeat elements was based on a custom *LOLA* database created using the UCSC Repeat Masker track (http://www.repeatmasker.org).

### Gene set overrepresentation and enrichment analysis

We used gene sets from the Molecular Signatures Database v. 6.2 (MSigDB) (84) to test for the overrepresentation of DEGs or genes associated with differential ATAC peaks, and the gene set enrichment analysis (GSEA) of DEGs. Overrepresentation analysis was performed with the help of R package *GeneOverlap* (v. 1. 14.0). GSEA was performed *with fgsea*package (v. 1.4.1) (85) using 10,000 permutations.

### Availability of data and code

All raw sequencing data from WGS, WGBS, ATAC-seq, ChIP-seq, RNA-seq and deep targeted resequencing were obtained from patient samples (therefore restricted) and are being uploaded to European Genome-Phenome Archive (EGA): EGA: EGAS00001003730. Processed sequencing data and microarray data are being uploaded to ArrayExpress database (E-MTAB-7184). All presented results were obtained with the use of published and publicly available software tools introduced in the Methods section.

## Data Availability

All raw sequencing data from WGS, WGBS, ATAC-seq, ChIP-seq, RNA-seq and deep targeted resequencing were obtained from patient samples (therefore restricted) and are being uploaded to European Genome-Phenome Archive (EGA): EGA: EGAS00001003730. Processed sequencing data and microarray data are being uploaded to the ArrayExpress database (E-MTAB-7184). All presented results were obtained with the use of published and publicly available software tools introduced in the Methods section.

## Acknowledgements

We acknowledge the Genomics and Proteomics Core Facility of the German Cancer Research Center (head Stephan Wiemann) for their assistance with the high-throughput sequencing, and the Omics and Data Management Core Facility (heads Jürgen Eils, Christian Lawerenz and Ivo Buchhalter) for the help with processing and management of the sequencing data. We are thankful to Christa Winkelmann (Institute of Pathology, Erlangen) for excellent performance of immunostainings, Cansu Cirzi for the help with ChIP-mentation libraries and Oliver Mücke for his help with MassARRAY. We thank ActiveMotif for providing a novel specific antibody against H3.3-G34W, as well as JPT (Pavlo Holenya and Ulf Reimer) for kindly providing a histone peptide array for antibody validation. We thank Stephan Pfister and David Jones for the permission to use PedGBM cohort, and Ana Banito for a thoughtful feedback about the manuscript.

## Author Contributions

C.P. and A.L. supervised the study. F.H., J.Z. and J.F. provided GCTB samples and performed their initial characterization. A.B., S.Ö., D.M., J.F., A. Jau., A.K., M.H., J.L., M. Schl., M.Schuh., D.V., F.G., S.H., V.H.N., S.L., J.H.P., Y.J.P., D.K., C.J., F.R. and D.W. performed experimental work. P.L., S.Ö., A.M., U.T., R.T., J.H., K.B., C.J., U.O. performed bioinformatical data analysis and interpretation. P.L., A.B., S.Ö., D.M., A.K., C.P. and A.L. wrote the manuscript.

## Competing interests’ statement

The authors declare no potential conflicts of interest.

Requests for materials and other correspondence should be directed to corresponding authors.

## References

1. Mohammad F, Helin K. Oncohistones: drivers of pediatric cancers. Genes Dev 2017;31(23–24):2313-24 doi 10.1101/gad.309013.117.

2. Nacev BA, Feng L, Bagert JD, Lemiesz AE, Gao J, Soshnev AA, et al. The expanding landscape of ‘oncohistone’ mutations in human cancers. Nature 2019 doi 10.1038/s41586-019-1038-1.

3. Bennett RL, Bele A, Small EC, Will CM, Nabet B, Oyer JA, et al. A Mutation in Histone H2B Represents a New Class of Oncogenic Driver. Cancer Discov 2019;9(10):1438–51 doi 10.1158/2159-8290.CD-19-0393.

4. Szenker E, Ray-Gallet D, Almouzni G. The double face of the histone variant H3.3. Cell Res 2011;21(3):421–34 doi 10.1038/cr.2011.14.

5. Goldberg AD, Banaszynski LA, Noh KM, Lewis PW, Elsaesser SJ, Stadler S, et al. Distinct factors control histone variant H3.3 localization at specific genomic regions. Cell 2010;140(5):678–91 doi 10.1016/j.cell.2010.01.003.

6. Voon HP, Wong LH. New players in heterochromatin silencing: histone variant H3.3 and the ATRX/DAXX chaperone. Nucleic acids research 2016;44(4):1496–501 doi 10.1093/nar/gkw012.

7. Banaszynski LA, Wen D, Dewell S, Whitcomb SJ, Lin M, Diaz N, et al. Hira-dependent histone H3.3 deposition facilitates PRC2 recruitment at developmental loci in ES cells. Cell 2013;155(1):107–20 doi 10.1016/j.cell.2013.08.061.

8. Bernstein BE, Mikkelsen TS, Xie X, Kamal M, Huebert DJ, Cuff J, et al. A bivalent chromatin structure marks key developmental genes in embryonic stem cells. Cell 2006;125(2):315–26 doi 10.1016/j.cell.2006.02.041.

9. Schwartzentruber J, Korshunov A, Liu XY, Jones DT, Pfaff E, Jacob K, et al. Driver mutations in histone H3.3 and chromatin remodelling genes in paediatric glioblastoma. Nature 2012;482(7384):226–31 doi 10.1038/nature10833.

10. Behjati S, Tarpey PS, Presneau N, Scheipl S, Pillay N, Van Loo P, et al. Distinct H3F3A and H3F3B driver mutations define chondroblastoma and giant cell tumor of bone. Nature genetics 2013;45(12):1479–82 doi 10.1038/ng.2814.

11. Fang D, Gan H, Lee JH, Han J, Wang Z, Riester SM, et al. The histone H3.3K36M mutation reprograms the epigenome of chondroblastomas. Science 2016;352(6291):1344–8 doi 10.1126/science.aae0065.

12. Chan KM, Fang D, Gan H, Hashizume R, Yu C, Schroeder M, et al. The histone H3.3K27M mutation in pediatric glioma reprograms H3K27 methylation and gene expression. Genes Dev 2013;27(9):985–90 doi 10.1101/gad.217778.113.

13. Lewis PW, Muller MM, Koletsky MS, Cordero F, Lin S, Banaszynski LA, et al. Inhibition of PRC2 Activity by a Gain-of-Function H3 Mutation Found in Pediatric Glioblastoma. Science 2013 doi 10.1126/science.1232245.

14. Bender S, Tang Y, Lindroth AM, Hovestadt V, Jones DT, Kool M, et al. Reduced H3K27me3 and DNA hypomethylation are major drivers of gene expression in K27M mutant pediatric high-grade gliomas. Cancer cell 2013;24(5):660–72 doi 10.1016/j.ccr.2013.10.006.

15. Lu C, Jain SU, Hoelper D, Bechet D, Molden RC, Ran L, et al. Histone H3K36 mutations promote sarcomagenesis through altered histone methylation landscape. Science 2016;352(6287):844–9 doi 10.1126/science.aac7272.

16. Shi L, Shi J, Shi X, Li W, Wen H. Histone H3.3 G34 Mutations Alter Histone H3K36 and H3K27 Methylation In Cis. J Mol Biol 2018 doi 10.1016/j.jmb.2018.04.014.

17. Amanatullah DF, Clark TR, Lopez MJ, Borys D, Tamurian RM. Giant cell tumor of bone. Orthopedics 2014;37(2):112–20 doi 10.3928/01477447-20140124-08.

18. Raskin KA, Schwab JH, Mankin HJ, Springfield DS, Hornicek FJ. Giant cell tumor of bone. J Am Acad Orthop Surg 2013;21(2):118–26 doi 10.5435/JAAOS-21-02-118.

19. Goldring SR, Roelke MS, Petrison KK, Bhan AK. Human giant cell tumors of bone identification and characterization of cell types. The Journal of clinical investigation 1987;79(2):483–91 doi 10.1172/JCI112838.

20. International Cancer Genome Consortium PedBrain Tumor P. Recurrent MET fusion genes represent a drug target in pediatric glioblastoma. Nat Med 2016;22(11):1314–20 doi 10.1038/nm.4204.

21. Roadmap Epigenomics C, Kundaje A, Meuleman W, Ernst J, Bilenky M, Yen A, et al. Integrative analysis of 111 reference human epigenomes. Nature 2015;518(7539):317–30 doi 10.1038/nature14248.

22. Zhou W, Dinh HQ, Ramjan Z, Weisenberger DJ, Nicolet CM, Shen H, et al. DNA methylation loss in late-replicating domains is linked to mitotic cell division. Nature genetics 2018;50(4):591–602 doi 10.1038/s41588-018-0073-4.

23. Moskovszky L, Szuhai K, Krenacs T, Hogendoorn PC, Szendroi M, Benassi MS, et al. Genomic instability in giant cell tumor of bone. A study of 52 cases using DNA ploidy, relocalization FISH, and array-CGH analysis. Genes Chromosomes Cancer 2009;48(6):468–79 doi 10.1002/gcc.20656.

24. Lim J, Park JH, Baude A, Yoo Y, Lee YK, Schmidt CR, et al. The histone variant H3.3 G34W substitution in giant cell tumor of the bone link chromatin and RNA processing. Sci Rep 2017;7(1):13459 doi 10.1038/s41598-017-13887-y.

25. Balke M. Denosumab treatment of giant cell tumour of bone. The Lancet Oncology 2013;14(9):801–2 doi 10.1016/S1470-2045(13)70291-2.

26. Boyle WJ, Simonet WS, Lacey DL. Osteoclast differentiation and activation. Nature 2003;423(6937):337–42 doi 10.1038/nature01658.

27. Kieslinger M, Folberth S, Dobreva G, Dorn T, Croci L, Erben R, et al. EBF2 regulates osteoblast-dependent differentiation of osteoclasts. Dev Cell 2005;9(6):757–67 doi 10.1016/j.devcel.2005.10.009.

28. Boyce BF, Xing L, Chen D. Osteoprotegerin, the bone protector, is a surprising target for beta-catenin signaling. Cell Metab 2005;2(6):344–5 doi 10.1016/j.cmet.2005.11.011.

29. Wolock SL, Krishnan I, Tenen DE, Matkins V, Camacho V, Patel S, et al. Mapping Distinct Bone Marrow Niche Populations and Their Differentiation Paths. Cell Rep 2019;28(2):302–11 e5 doi 10.1016/j.celrep.2019.06.031.

30. Zardo G, Tiirikainen MI, Hong C, Misra A, Feuerstein BG, Volik S, et al. Integrated genomic and epigenomic analyses pinpoint biallelic gene inactivation in tumors. Nature genetics 2002;32(3):453–8 doi 10.1038/ng1007.

31. Shapira SN, Lim HW, Rajakumari S, Sakers AP, Ishibashi J, Harms MJ, et al. EBF2 transcriptionally regulates brown adipogenesis via the histone reader DPF3 and the BAF chromatin remodeling complex. Genes Dev 2017;31(7):660–73 doi 10.1101/gad.294405.116.

32. Voon HP, Hughes JR, Rode C, De La Rosa-Velazquez IA, Jenuwein T, Feil R, et al. ATRX Plays a Key Role in Maintaining Silencing at Interstitial Heterochromatic Loci and Imprinted Genes. Cell Rep 2015;11(3):405–18 doi 10.1016/j.celrep.2015.03.036.

33. Harutyunyan AS, Krug B, Chen H, Papillon-Cavanagh S, Zeinieh M, De Jay N, et al. H3K27M induces defective chromatin spread of PRC2-mediated repressive H3K27me2/me3 and is essential for glioma tumorigenesis. Nat Commun 2019;10(1):1262 doi 10.1038/s41467-019-09140-x.

34. Hardouin SN, Guo R, Romeo PH, Nagy A, Aubin JE. Impaired mesenchymal stem cell differentiation and osteoclastogenesis in mice deficient for Igf2-P2 transcripts. Development 2011;138(2):203–13 doi 10.1242/dev.054916.

35. Xu JC, Wu GH, Zhou LL, Yang XJ, Liu JT. Leptin improves osteoblast differentiation of human bone marrow stroma stem cells. Eur Rev Med Pharmacol Sci 2016;20(16):3507–13.

36. Huang L, Teng XY, Cheng YY, Lee KM, Kumta SM. Expression of preosteoblast markers and Cbfa-1 and Osterix gene transcripts in stromal tumour cells of giant cell tumour of bone. Bone 2004;34(3):393–401 doi 10.1016/j.bone.2003.10.013.

37. Lau CP, Kwok JS, Tsui JC, Huang L, Yang KY, Tsui SK, et al. Genome-Wide Transcriptome Profiling of the Neoplastic Giant Cell Tumor of Bone Stromal Cells by RNA Sequencing. J Cell Biochem 2017;118(6):1349–60 doi 10.1002/jcb.25792.

38. Zhang Y, Shan C-M, Wang J, Bao K, Tong L, Jia S. Molecular basis for the role of oncogenic histone mutations in modulating H3K36 methylation. Scientific Reports 2017;7:43906 doi 10.1038/srep43906.

39. Sturm D, Witt H, Hovestadt V, Khuong-Quang DA, Jones DT, Konermann C, et al. Hotspot Mutations in H3F3A and IDH1 Define Distinct Epigenetic and Biological Subgroups of Glioblastoma. Cancer cell 2012;22(4):425–37 doi 10.1016/j.ccr.2012.08.024.

40. de la Rica L, Rodriguez-Ubreva J, Garcia M, Islam AB, Urquiza JM, Hernando H, et al. PU.1 target genes undergo Tet2-coupled demethylation and DNMT3b-mediated methylation in monocyte-to-osteoclast differentiation. Genome Biol 2013;14(9):R99 doi 10.1186/gb-2013-14-9-r99.

41. Rauch A, Haakonsson AK, Madsen JGS, Larsen M, Forss I, Madsen MR, et al. Osteogenesis depends on commissioning of a network of stem cell transcription factors that act as repressors of adipogenesis. Nature genetics 2019 doi 10.1038/s41588-019-0359-1.

42. Lau YS, Sabokbar A, Gibbons CL, Giele H, Athanasou N. Phenotypic and molecular studies of giant-cell tumors of bone and soft tissue. Hum Pathol 2005;36(9):945–54 doi 10.1016/j.humpath.2005.07.005.

43. Murata A, Fujita T, Kawahara N, Tsuchiya H, Tomita K. Osteoblast lineage properties in giant cell tumors of bone. J Orthop Sci 2005;10(6):581–8 doi 10.1007/s00776-005-0946-0.

44. Chiellini C, Grenningloh G, Cochet O, Scheideler M, Trajanoski Z, Ailhaud G, et al. Stathmin-like 2, a developmentally-associated neuronal marker, is expressed and modulated during osteogenesis of human mesenchymal stem cells. Biochem Biophys Res Commun 2008;374(1):64–8 doi 10.1016/j.bbrc.2008.06.121.

45. Liu L, Aleksandrowicz E, Fan P, Schonsiegel F, Zhang Y, Sahr H, et al. Enrichment of c-Met+ tumorigenic stromal cells of giant cell tumor of bone and targeting by cabozantinib. Cell Death Dis 2014;5:e1471 doi 10.1038/cddis.2014.440.

46. Wulling M, Delling G, Kaiser E. The origin of the neoplastic stromal cell in giant cell tumor of bone. Hum Pathol 2003;34(10):983–93 doi 10.1053/s0046-8177(03)00413-1.

47. Baubec T, Colombo DF, Wirbelauer C, Schmidt J, Burger L, Krebs AR, et al. Genomic profiling of DNA methyltransferases reveals a role for DNMT3B in genic methylation. Nature 2015;520(7546):243–7 doi 10.1038/nature14176.

48. Dhayalan A, Rajavelu A, Rathert P, Tamas R, Jurkowska RZ, Ragozin S, et al. The Dnmt3a PWWP domain reads histone 3 lysine 36 trimethylation and guides DNA methylation. J Biol Chem 2010;285(34):26114–20 doi 10.1074/jbc.M109.089433.

49. Souren NY, Gerdes LA, Kumpfel T, Lutsik P, Klopstock T, Hohlfeld R, et al. Mitochondrial DNA Variation and Heteroplasmy in Monozygotic Twins Clinically Discordant for Multiple Sclerosis. Hum Mutat 2016;37(8):765–75 doi 10.1002/humu.23003.

50. Li H, Durbin R. Fast and accurate short read alignment with Burrows-Wheeler transform. Bioinformatics 2009;25(14):1754–60 doi 10.1093/bioinformatics/btp324.

51. Gerstung M, Beisel C, Rechsteiner M, Wild P, Schraml P, Moch H, et al. Reliable detection of subclonal single-nucleotide variants in tumour cell populations. Nat Commun 2012;3:811 doi 10.1038/ncomms1814.

52. Geigl JB, Uhrig S, Speicher MR. Multiplex-fluorescence in situ hybridization for chromosome karyotyping. Nat Protoc 2006;1(3):1172–84 doi 10.1038/nprot.2006.160.

53. Wang K, Li M, Hakonarson H. ANNOVAR: functional annotation of genetic variants from high-throughput sequencing data. Nucleic acids research 2010;38(16):e164 doi 10.1093/nar/gkq603.

54. Tischler G, Leonard S. biobambam: tools for read pair collation based algorithms on BAM files. Source Code for Biology and Medicine 2014;9(1):13 doi 10.1186/1751-0473-9-13.

55. Jones DT, Hutter B, Jager N, Korshunov A, Kool M, Warnatz HJ, et al. Recurrent somatic alterations of FGFR1 and NTRK2 in pilocytic astrocytoma. Nature genetics 2013;45(8):927–32 doi 10.1038/ng.2682.

56. Rimmer A, Phan H, Mathieson I, Iqbal Z, Twigg SRF, Consortium WGS, et al. Integrating mapping-, assembly- and haplotype-based approaches for calling variants in clinical sequencing applications. Nature genetics 2014;46(8):912–8 doi 10.1038/ng.3036.

57. Sahm F, Toprak UH, Hubschmann D, Kleinheinz K, Buchhalter I, Sill M, et al. Meningiomas induced by low-dose radiation carry structural variants of NF2 and a distinct mutational signature. Acta Neuropathol 2017; 134(1):155–8 doi 10.1007/s00401-017-1715-9.

58. Kleinheinz K, Bludau I, Huebschmann D, Heinold M, Kensche P, Gu Z, et al. ACEseq - allele specific copy number estimation from whole genome sequencing. bioRxiv 2017 doi 10.1101/210807.

59. Buenrostro JD, Giresi PG, Zaba LC, Chang HY, Greenleaf WJ. Transposition of native chromatin for fast and sensitive epigenomic profiling of open chromatin, DNA-binding proteins and nucleosome position. Nat Methods 2013;10(12):1213–8 doi 10.1038/nmeth.2688.

60. Martin M. Cutadapt removes adapter sequences from high-throughput sequencing reads. 2011 2011;17(1):3 doi 10.14806/ej.17.1.200.

61. Langmead B, Salzberg SL. Fast gapped-read alignment with Bowtie 2. Nat Methods 2012;9(4):357–9 doi 10.1038/nmeth.1923.

62. Ramirez F, Dundar F, Diehl S, Gruning BA, Manke T. deepTools: a flexible platform for exploring deep-sequencing data. Nucleic acids research 2014;42(Web Server issue):W187–91 doi 10.1093/nar/gku365.

63. Zhang Y, Liu T, Meyer CA, Eeckhoute J, Johnson DS, Bernstein BE, et al. Model-based analysis of ChIP-Seq (MACS). Genome Biol 2008;9(9):R137 doi 10.1186/gb-2008-9-9-r137.

64. McCarthy DJ, Chen Y, Smyth GK. Differential expression analysis of multifactor RNA-Seq experiments with respect to biological variation. Nucleic acids research 2012;40(10):4288–97 doi 10.1093/nar/gks042.

65. Zhu LJ. Integrative analysis of ChIP-chip and ChIP-seq dataset. Methods Mol Biol 2013;1067:105–24 doi 10.1007/978-1-62703-607-8_8.

66. Heinz S, Benner C, Spann N, Bertolino E, Lin YC, Laslo P, et al. Simple combinations of lineage-determining transcription factors prime cis-regulatory elements required for macrophage and B cell identities. Mol Cell 2010;38(4):576–89 doi 10.1016/j.molcel.2010.05.004.

67. Schmidl C, Rendeiro AF, Sheffield NC, Bock C. ChIPmentation: fast, robust, low-input ChIP-seq for histones and transcription factors. Nat Methods 2015;12(10):963–5 doi 10.1038/nmeth.3542.

68. Ernst J, Kellis M. Chromatin-state discovery and genome annotation with ChromHMM. Nat Protoc 2017;12(12):2478–92 doi 10.1038/nprot.2017.124.

69. Bolger AM, Lohse M, Usadel B. Trimmomatic: a flexible trimmer for Illumina sequence data. Bioinformatics 2014;30(15):2114–20 doi 10.1093/bioinformatics/btu170.

70. Hansen KD, Langmead B, Irizarry RA. BSmooth: from whole genome bisulfite sequencing reads to differentially methylated regions. Genome Biol 2012;13(10):R83 doi 10.1186/gb-2012-13-10-r83.

71. Park Y, Wu H. Differential methylation analysis for BS-seq data under general experimental design. Bioinformatics 2016;32(10):1446–53 doi 10.1093/bioinformatics/btw026.

72. Assenov Y, Muller F, Lutsik P, Walter J, Lengauer T, Bock C. Comprehensive analysis of DNA methylation data with RnBeads. Nat Methods 2014; 11(11):1138–40 doi 10.1038/nmeth.3115.

73. Weischenfeldt J, Simon R, Feuerbach L, Schlangen K, Weichenhan D, Minner S, et al. Integrative genomic analyses reveal an androgen-driven somatic alteration landscape in early-onset prostate cancer. Cancer cell 2013;23(2):159–70 doi 10.1016/j.ccr.2013.01.002.

74. Kim D, Langmead B, Salzberg SL. HISAT: a fast spliced aligner with low memory requirements. Nat Methods 2015;12(4):357–60 doi 10.1038/nmeth.3317.

75. Pertea M, Pertea GM, Antonescu CM, Chang TC, Mendell JT, Salzberg SL. StringTie enables improved reconstruction of a transcriptome from RNA-seq reads. Nat Biotechnol 2015;33(3):290–5 doi 10.1038/nbt.3122.

76. Love MI, Huber W, Anders S. Moderated estimation of fold change and dispersion for RNA-seq data with DESeq2. Genome Biol 2014;15(12):550 doi 10.1186/s13059-014-0550-8.

77. Bray NL, Pimentel H, Melsted P, Pachter L. Near-optimal probabilistic RNA-seq quantification. Nat Biotechnol 2016;34(5):525–7 doi 10.1038/nbt.3519.

78. Court F, Arnaud P. An annotated list of bivalent chromatin regions in human ES cells: a new tool for cancer epigenetic research. Oncotarget 2017;8(3):4110–24 doi 10.18632/oncotarget.13746.

79. Bracken AP, Dietrich N, Pasini D, Hansen KH, Helin K. Genome-wide mapping of Polycomb target genes unravels their roles in cell fate transitions. Genes Dev 2006;20(9): 1123-36 doi 10.1101/gad.381706.

80. Skaar DA, Li Y, Bernal AJ, Hoyo C, Murphy SK, Jirtle RL. The human imprintome: regulatory mechanisms, methods of ascertainment, and roles in disease susceptibility. ILAR J 2012;53(3–4):341-58 doi 10.1093/ilar.53.3-4.341.

81. Hansen RS, Thomas S, Sandstrom R, Canfield TK, Thurman RE, Weaver M, et al. Sequencing newly replicated DNA reveals widespread plasticity in human replication timing. Proceedings of the National Academy of Sciences of the United States of America 2010;107(1):139–44 doi 10.1073/pnas.0912402107.

82. Zynda GJ, Song J, Concia L, Wear EE, Hanley-Bowdoin L, Thompson WF, et al. Repliscan: a tool for classifying replication timing regions. BMC Bioinformatics 2017;18(1):362 doi 10.1186/s12859-017-1774-x.

83. Sheffield NC, Bock C. LOLA: enrichment analysis for genomic region sets and regulatory elements in R and Bioconductor. Bioinformatics 2016;32(4):587–9 doi 10.1093/bioinformatics/btv612.

84. Subramanian A, Tamayo P, Mootha VK, Mukherjee S, Ebert BL, Gillette MA, et al. Gene set enrichment analysis: a knowledge-based approach for interpreting genome-wide expression profiles. Proceedings of the National Academy of Sciences of the United States of America 2005;102(43):15545–50 doi 10.1073/pnas.0506580102.

85. Sergushichev A. An algorithm for fast preranked gene set enrichment analysis using cumulative statistic calculation. bioRxiv 2016 doi 10.1101/060012.

